# Treatment escalation after clinically silent MRI lesions in relapsing-remitting multiple sclerosis

**DOI:** 10.64898/2026.03.09.26347918

**Authors:** Cyrus Daruwalla, Courtney Kremler, Francesco Patti, Serkan Ozakbas, Cavit Boz, Jeannette Lechner-Scott, Andrea Surcinelli, Matteo Foschi, Samia J Khoury, Helmut Butzkueven, Anneke van der Walt, Zuzana Rous, Mario Habek, Jose E Meca-Lallana, Gabriel Valero Lopez, Raed Alroughani, Yolanda Blanco, Guy Laureys, Olga Skibina, Katherine Buzzard, Orla Gray, Pamela McCombe, Davide Maimone, Pierre Duquette, Marc Girard, Alexandre Prat, Jose Luis Sanchez-Menoyo, Vincent van Pesch, Aysun Soysal, Maria Pia Amato, Francois Grand’Maison, Judith Wilton, Bart Van Wijmeersch, Oliver Gerlach, Alessandra Lugaresi, Valentina Tomassini, Giovanna De Luca, Bruce Taylor, Yi Chao Foong, Nevin John, Simón Cárdenas-Robledo, Suzanne Hodgkinson, Richard Macdonell, Barbara Willekens, Justin Garber, Emanuele D’Amico, Murat Terzi, Jana Houskova, Emmanuelle Lapointe, Abdullah Al-Asmi, Michael Barnett, Ivana Stetkarova, Riadh Gouider, Saloua Mrabet, Tunde Csepany, Chris McGuigan, Katrin Gross-Paju, Jens Kuhle, Tamara Castillo-Triviño, Simon Kerrigan, Aviva M Tolkovsky, Mark Wardle, Nick G Cunniffe, Izanne Roos, Paul Madley-Dowd, Alasdair Coles, Tomas Kalincik, Jonathan A C Sterne, J William L Brown, the MSBase study group

## Abstract

Clinically silent MRI lesions occur frequently in people with relapsing-remitting multiple sclerosis (RRMS) despite disease modifying therapy (DMT). Guidelines only recommend DMT escalation after multiple silent lesions, and adherence is variable. We explored outcomes and the effect of treatment escalation following single and multiple on-treatment silent lesions.

This cohort study and emulated target trial used MSBase registry data from 99 clinics in 26 countries between 2007 and 2025. Clinically stable participants receiving any DMT for RRMS with silent lesions versus without silent lesions were compared. Among participants with silent lesions while taking platform or moderate-efficacy DMTs, outcomes following treatment escalation within 6 months versus no treatment escalation (unless a post-MRI clinical event occurred) were compared. The primary outcome was an MS relapse, and the secondary outcome was 6-month confirmed disability worsening.

A total of 10,232 participants met inclusion criteria (71.7% female, mean age 41 [SD 11]). The 2-year cumulative incidence of relapse was 27.8% (95% CI 25.7%-29.9%) in participants with silent lesions versus 14.3% (95% CI 13.5%-15.2%) without (adjusted hazard ratio [aHR] 1.76 [95% CI 1.57-1.97]). The 2-year cumulative incidence of disability worsening was 13.8% (95% CI 12.2%-15.5%) in participants with silent lesions versus 11.4% (95% CI 10.7%-12.2%) without (aHR 1.38 [95% CI 1.18-1.62]). Rates of relapse and disability worsening were higher following single and multiple silent lesions versus no silent lesions.

The emulated trial included 2,264 participants with ≥1 silent lesion on platform or moderate efficacy DMTs, 286 of whom escalated DMT within 6 months following silent lesions. The 4-year cumulative incidence of relapse was lower following treatment escalation (16.8% [95% CI 12.4%-23.4%]) versus continuation (38.9% [95% CI 35.8%-42.1%]), aHR 0.34 (95% CI 0.23-0.47), with similar aHRs following single and multiple silent lesions. The 4-year cumulative incidence of disability worsening was similar following treatment escalation (16.0% [95% CI 10.8%-22.2%]) versus continuation (17.7% [95% CI 15.3%-20.1%]), aHR 0.89 (95% CI 0.56-1.33).

People with RRMS with single or multiple on-treatment silent MRI lesions have higher subsequent risks of relapse and disability worsening than people without silent lesions. DMT escalation mitigates the relapse risk, though disability worsening continues at a similar rate over 4 years. Contrary to guidelines, DMT escalation should be considered after single or multiple silent lesions.

## Introduction

Multiple sclerosis (MS) is among the leading causes of disability in young adults.^1^ Early treatment with high-efficacy disease modifying therapies (DMTs) attenuates disability worsening in people with relapsing-remitting (RR) MS,^2–5^ but high-efficacy DMTs often carry significant risks, administration and monitoring burdens, and costs. Initial treatment with platform and moderate efficacy DMTs therefore remains common.^6^ Clinical guidelines recommend annual on-treatment MRI head scans^7^ and DMT escalation in response to clinical relapses or multiple clinically silent lesions (CSLs)^8–10^ – new, expanding, or contrast-enhancing MRI lesions in the absence of relapses or disability worsening – which emerge around nine times more frequently than clinical relapses.^11^

Adherence to guidelines is variable. Most participants in cohort studies using data from clinical settings did not escalate DMT following multiple CSLs,^12,13^ and a recent worldwide survey found that <60% of clinicians would escalate DMT following two new MRI lesions accompanied by new neurological examination findings.^6^ This reflects conflicting evidence. Previous studies found either no association between CSLs and adverse outcomes^14–16^ or associations only following multiple CSLs versus no CSLs.^13,17–19^ Subsequent DMT escalation was associated with improved outcomes only following multiple CSLs.^13^ Hence, while 40% of scans revealing breakthrough disease activity demonstrate a single CSL,^12^ this is sometimes termed ‘minimal evidence of disease activity’ (MEDA)^17,18,20^ and treatment escalation is not recommended in current clinical guidelines.^8–10^

Using a large international registry, we compared relapse and disability worsening outcomes in people on DMTs for RRMS with single and multiple CSLs compared to no CSLs. We then emulated a target trial^21–24^ examining the effect of post-CSL DMT escalation versus no escalation on the same outcomes.

## Participants and methods

### Participants

This cohort study and emulated target trial used data from the international MSBase neuroimmunology registry,^25^ approved by the Melbourne Health Human Research Ethics Committee and ethics committees in participating centres (or exemptions granted according to local regulations; WHO-ICTRP-ID: ACTRN12605000455662). All participants provided consent as per local regulations. MSBase contains patient-level data collected during routine clinical care in 203 clinics in 45 countries. Data have been entered prospectively since July 1, 2004, but retrospective data can also be entered. Data were extracted on 1^st^ April 2025.

#### Eligibility criteria

Eligibility criteria (detailed in the Supplementary Methods) were designed to identify contemporary, clinically stable participants receiving DMTs for adult-onset RRMS, with MRI scans at approximately annual intervals.^7^ Hence, eligible participants had at least one pair of MRI brain scans: an “index MRI” performed after 2006, and a “comparator MRI” performed 6 to 18 months earlier. Eligible scan pairs required treatment with a single DMT (any DMT for prognosis analyses; platform or moderate-efficacy DMTs for emulated trials) throughout the interscan interval. To account for therapeutic lag,^26^ and mirror current guidelines requiring a “re-baseline” scan,^7^ ≥6 months of treatment were required before the comparator MRI (or ≥18 months for induction treatments given over 12 [alemtuzumab] or 13 months [cladribine]). To satisfy the positivity assumption, MRIs performed before availability of higher-efficacy DMTs in each participant’s treating centre were excluded. To ensure clinical stability, scan pairs were excluded if there were relapses or Expanded Disability Status Scale (EDSS) score increases (from a baseline ≤12 months before the comparator MRI) in the interscan interval. Participants required at least one follow-up visit for relapse analyses, or at least two follow-up EDSS scores ≥6 months apart for 6-month confirmed disability worsening analyses. Participants were excluded if no eligible participants in their country had the outcome of interest.

### Procedures

#### MRI scans

MRI scans were performed during routine clinical care, typically including T2-weighted or fluid-attenuated inversion recovery and T1-weighted (with or without gadolinium contrast) sequences at 1.5 or 3T field strength, with 1–2 mm isometric or non-isometric voxel size. The number of new or enlarging T2 and enhancing T1 lesions was recorded in accordance with local clinical practice, including radiologist review. Where data were missing, primary source verification was sought from local investigators. The index MRI was deemed to contain CSLs if it contained ≥1 new or enlarging T2-hyperintense lesion compared to the comparator MRI, or ≥1 contrast-enhancing lesion.

#### Disease modifying therapy efficacy groups

DMTs were categorised according to their efficacy in preventing relapses into platform (glatiramer acetate, interferon-beta, and teriflunomide), moderate-efficacy (cladribine, dimethyl fumarate, diroximel fumarate, fingolimod, ozanimod, ponesimod, and siponimod) and high-efficacy (alemtuzumab, autologous haematopoietic stem cell transplantation, natalizumab, ocrelizumab, ofatumumab, and rituximab) groups.^27^

#### Outcomes

The primary outcome was an MS relapse (a symptomatic CNS demyelinating event despite no fever or infection, ≥30 days after the previous relapse, lasting ≥24 hours). The secondary outcome was 6-month confirmed disability worsening (CDW; an EDSS score increase of ≥1.5 if the baseline was 0, ≥1.0 if the baseline was between 1.0 and 5.5, and ≥0.5 if the baseline was ≥6.0, confirmed ≥6 months later).^28^ Exploratory outcomes were subtypes of disability worsening: relapse-associated worsening (RAW) and progression independent of relapse activity (PIRA).^29^ Standard outcome definitions were used (detailed in the Supplementary Methods).

### Statistical analysis

#### Prognosis following clinically silent lesions

Starting from each eligible participant’s earliest index MRI, hazard ratios comparing participants with versus without CSLs were estimated using Cox models adjusted for potential baseline confounders: sex (self-reported), age, disease duration, calendar year, DMT efficacy group, EDSS score, and number of relapses in the prior 2 years (including polynomial terms for continuous variables as appropriate), and stratified by country (grouping countries with fewer than forty participants). The cumulative incidence of outcomes was derived from unadjusted Kaplan-Meier estimates. Participants with no CSLs were also compared to participants with 1 or ≥2 CSLs, and with contrast-enhancing and non-enhancing CSLs (among participants who received contrast on their index MRI). Subgroup analyses explored variation in associations with baseline DMT efficacy and disease duration. Sensitivity analyses: (1) replaced country stratification with adjustment for country health expenditure; (2) were conducted with and without adjusting models for the index MRI’s total number of T2 lesions (old and new), among participants with these data recorded; and (3) analysed unconfirmed disability worsening. Further detail is provided in the Supplementary Methods.

#### Emulated trial of DMT escalation following clinically silent lesions

A target trial emulation framework^21–23^ was used to specify a hypothetical randomised “target trial” and its emulation using non-randomised registry data (Supplementary Table 1). Among participants who fulfilled eligibility criteria (outlined above under “Participants”), their earliest MRI scan demonstrating ≥1 CSL while taking a platform or moderate-efficacy DMT was selected. Starting each week for 6 months following this MRI, the following non-blinded treatment strategies were compared among participants who had not escalated DMT by the end of the previous week:

1. DMT escalation
2. Control: no DMT escalation unless a clinical event occurs during follow-up, in which case DMT escalation must occur during the subsequent 6 months. Specifying DMT escalation following clinical events ensured that control participants did not act as proxies for a conservative approach to clinical events. For relapse outcome analyses, DMT escalation was required in response to disability worsening (but not relapses since these were the outcome) and vice versa.

Each week, participants who escalated DMT during the week were assigned to the DMT escalation group and all other remaining participants were assigned to the control group. This sequential approach^21,23,24,30^ aligned eligibility, treatment assignment, and the start of follow-up (baseline) for each of the 26 comparisons.

Hazard ratios comparing relapse and CDW outcomes were estimated using pooled logistic regression models adjusted for baseline confounders: sex (self-reported), age, disease duration, calendar year, DMT efficacy group, EDSS score, number of relapses in the prior 2 years, CSL number, contrast-enhancement, the number of post-CSL weeks, and country of care (a sensitivity analysis instead adjusted for country health expenditure, as did subgroup analyses); polynomial terms for continuous variables were used as appropriate. Since participants in the control arm in earlier weeks could escalate DMT in later weeks, we estimated the per protocol effect by censoring their follow-up if they escalated DMT despite not having a clinical event. Follow-up in control participants was also censored if they had not escalated DMT by six months after a clinical event. To mitigate selection bias introduced by this censoring, outcome models were weighted by the cumulative inverse probability of remaining uncensored, derived using logistic regression models adjusted for post-baseline values of confounders (listed above, plus the presence or absence of new lesions on follow-up MRI scans). Cumulative incidences were estimated using G-computation.^31^ Follow up continued until the earliest of outcome occurrence, deviation from the assigned treatment strategy, the last clinic visit, or 4 years. Due to the sequential emulation design, participants could contribute to the control group in multiple weeks, so participant-level bootstrapping with 1,000 random samples was used to derive 95% CIs. Sensitivity analyses: (i) analysed unconfirmed disability worsening; and (ii) compared DMT escalation to alternative control treatment strategies: (a) without requiring DMT escalation following clinical events; and (b) additionally without requiring unchanged DMT if there is no clinical event (analogous to the intention-to-treat effect). A detailed protocol is provided in the Supplementary Methods. Analyses were conducted in R version 4.3.1.

## Results

### Prognosis following clinically silent lesions

Of 90,998 MSBase participants with adult-onset RRMS, 10,232 participants from 99 clinics in 26 countries (Supplementary Table 2) were eligible (Fig. 1). The annualised relapse rate was similar in included (median 0.24 [IQR 0.14-0.37]) and excluded (0.25 [IQR 0.12-0.46]) participants. On their first eligible index MRI, 8,301 (81.1%) had no CSLs, 939 (9.2%) had a single CSL and 992 (9.7%) had multiple CSLs. Baseline characteristics and DMTs are in Supplementary Tables 3 and 4. At baseline, a higher proportion of participants with CSLs were on platform DMTs (1,247 of 1,931 [64.6%]) than participants without CSLs (3,301 of 8,301 [39.8%]). Post-baseline DMT escalation was more common in participants with (563 [29.1%]) versus without (675 [8.1%]) CSLs. There were no missing data on variables used to adjust for confounding.

**Figure 1.**
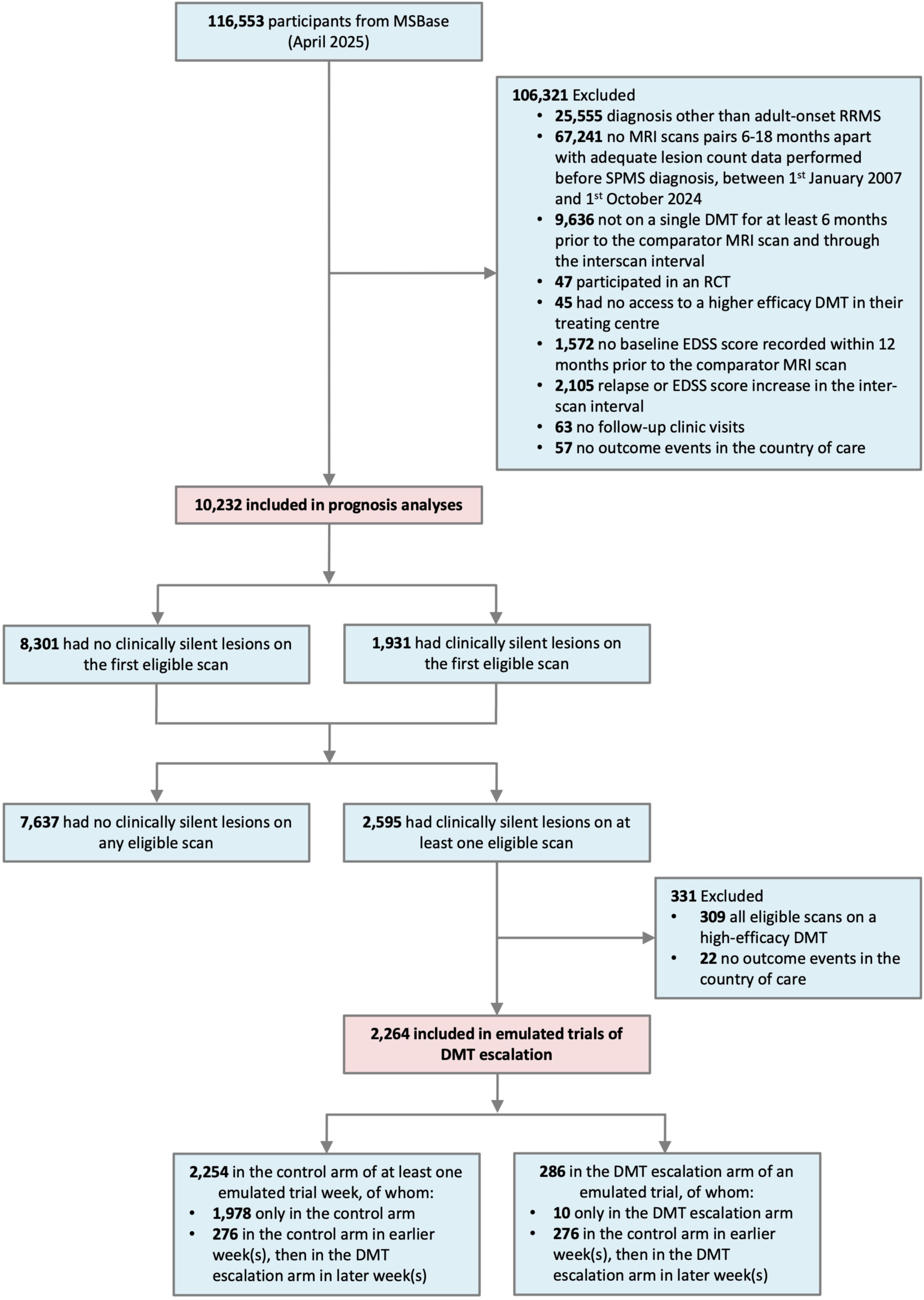
Inclusion flow diagram for analyses of relapse outcomes Abbreviations: DMT, disease modifying therapy; EDSS, Expanded Disability Status Scale; RCT, Randomised Controlled Trial; RRMS, relapsing-remitting multiple sclerosis; SPMS, secondary progressive multiple sclerosis.

Estimated hazard ratios from adjusted and unadjusted Cox models are in Supplementary Tables 5 and 6. During 2 years of follow-up, the cumulative incidence of relapse was 14.3% (95% CI 13.5%-15.2%) in participants without CSLs at baseline; 27.8% (95% CI 25.7%-29.9%) in participants with ≥1 CSL (adjusted hazard ratio [aHR] 1.76 [95% CI 1.57-1.97]); 25.1% (95% CI 22.3%-28.2%) in participants with 1 CSL (aHR 1.59 [95% CI 1.37-1.85]); and 30.3% (95% CI 27.4%-33.4%) in participants with multiple CSLs (aHR 1.94 [95% CI 1.68-2.24]; Fig. 2A). Both contrast-enhancing and non-enhancing CSLs were associated with increased rates of relapse (Supplementary Fig. 1A). Associations of CSLs with relapse were similar in subgroups defined by disease duration, and platform or moderate-efficacy baseline DMTs, but imprecisely estimated for high-efficacy DMTs (Supplementary Fig. 1A).

**Figure 2.**
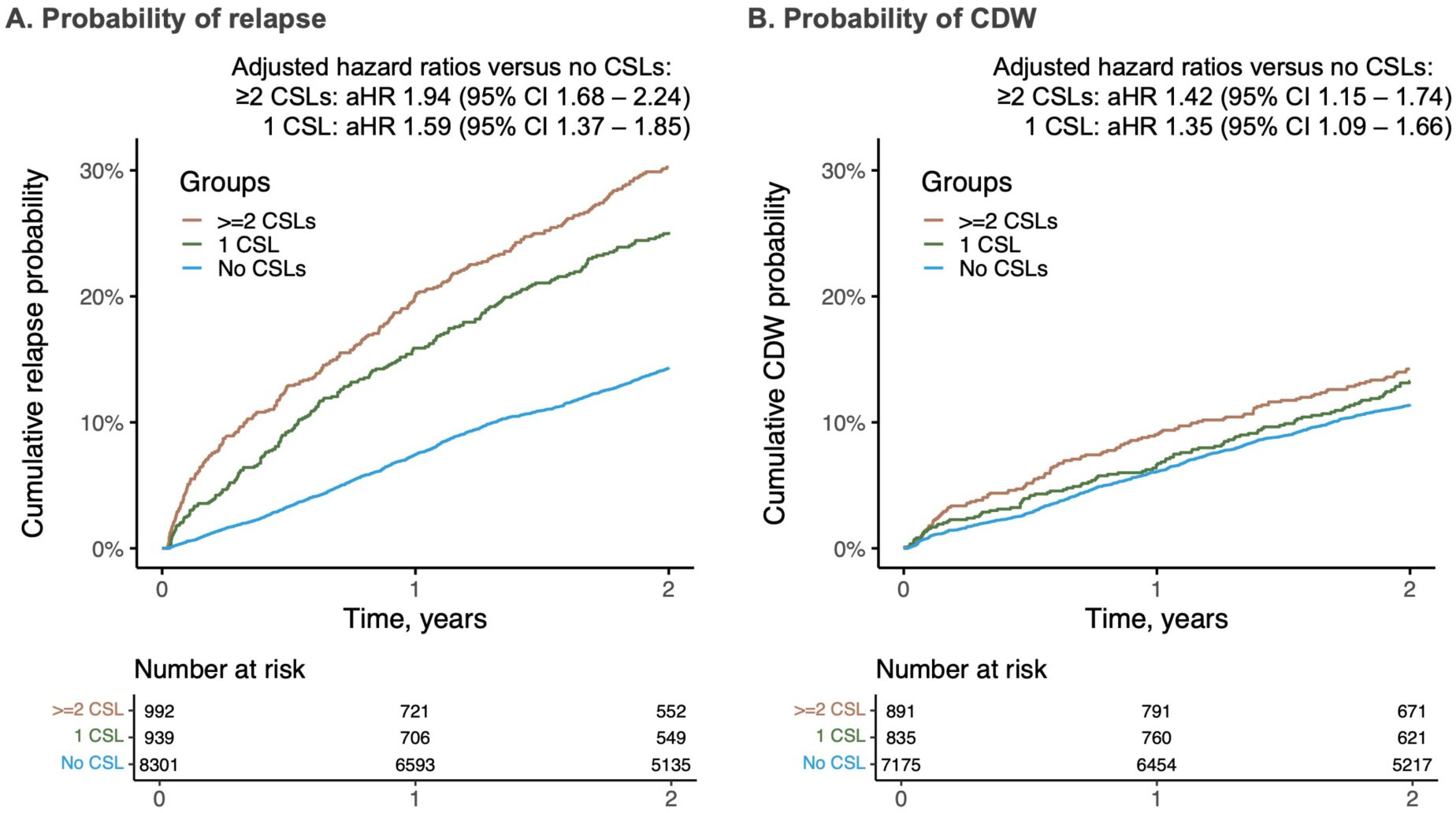
Probability of outcomes following single and multiple CSLs compared to no CSLs Kaplan-Meier curves show crude data for (A) relapse and (B) CDW outcomes. Adjusted hazard ratios compare outcomes in participants with 1 CSL (green) and ≥2 CSLs (brown) to no CSLs (blue) using Cox proportional hazards models adjusted for baseline confounders. Abbreviations: aHR, adjusted hazard ratio; CDW, confirmed disability worsening; CSL, clinically silent lesion.

Among 8,901 participants eligible for CDW assessment (Supplementary Fig. 2), the cumulative incidence of CDW was 11.4% (95% CI 10.7%-12.2%) in participants without CSLs; 13.8% (95% CI 12.2%-15.5%) in participants with ≥1 CSL (aHR 1.38 [95% CI 1.18-1.62]); 13.3% (95% CI 11.1%-15.8%) in participants with 1 CSL (aHR 1.35 [95% CI 1.09-1.66]); and 14.2% (95% CI 12.1%-16.8%) in participants with multiple CSLs (aHR 1.42 [95% CI 1.15-1.74]; Fig. 2B). Non-enhancing CSLs were associated with higher rates of CDW, but the association was imprecisely estimated for contrast-enhancing CSLs (Supplementary Fig. 1B). Associations of CSLs with CDW were similar in subgroups defined by baseline DMT efficacy and disease duration (Supplementary Fig. 1B). The cumulative incidence of RAW was 2.6% (95% CI 2.2%-3.0%) in participants without CSLs, and 5.1% (95% CI 4.1%-6.2%) in participants with CSLs (aHR 1.65 [95% CI 1.25-2.19]). The cumulative incidence of PIRA was 9.0% (95% CI 8.3%-9.7%) in participants without CSLs, and 9.0% (95% CI 7.7%-10.5%) in participants with CSLs (aHR 1.24 [95% CI 1.03-1.51]).

In sensitivity analyses comparing CSLs to no CSLs (Supplementary Table 7), Cox models adjusted for country health expenditure had similar adjusted hazard ratios to models stratified by country. Adjusted hazard ratios were also similar regardless of adjustment for total index-MRI T2 lesion count. Adjusted hazard ratios for unconfirmed disability worsening were similar to the main CDW outcome.

### Emulated trial of DMT escalation following clinically silent lesions

Of 2,264 participants eligible for the emulated trial assessing relapse outcomes, 286 escalated DMT within 6 months (Fig. 1). Table 1 and Supplementary Table 8 show participant characteristics and countries of care, respectively. Participants who escalated DMT within 6 months were younger and had a shorter disease duration; later year of inclusion; different distribution of countries; more CSLs; and more contrast-enhancing CSLs compared with all eligible participants; follow-up duration and frequency were similar. There were no missing data on variables used to adjust for confounding. Participants were included in a median of 26 emulated trials.

**Table 1.** Characteristics of participants included in the emulated trials of DMT escalation after clinically silent lesions.

|  | Relapse analysis |  |  | Confirmed disability worsening analysis |  |  |
| --- | --- | --- | --- | --- | --- | --- |
|  | All participants<sup>a</sup><br>(n=2264) | Participants who<br>escalated DMT within<br>6 months (n=286) | SMD | All participants<sup>a</sup><br>(n=2016) | Participants who<br>escalated DMT within<br>6 months (n=248) | SMD |
| **Sex, number (%)** |  |  | 0.027 |  |  | 0.021 |
| **Female** | 1587 (70.1%) | 204 (71.3%) |  | 1420 (70.4%) | 177 (71.4%) |  |
| **Male** | 677 (29.9%) | 82 (28.7%) |  | 596 (29.6%) | 71 (28.6%) |  |
| **Age, mean (SD)** | 39.52 (10.48) | 36.82 (10.03) | 0.263 | 39.29 (10.33) | 36.84 (10.17) | 0.239 |
| **Disease duration, median (IQR)** | 7.33 (4.15, 12.52) | 5.83 (3.59, 10.58) | 0.233 | 7.28 (4.15, 12.41) | 5.93 (3.57, 10.65) | 0.215 |
| **Year, median (IQR)** | 2017 (2014, 2020) | 2019 (2016, 2021) | 0.385 | 2017 (2014, 2020) | 2018 (2016, 2021) | 0.409 |
| **EDSS score, median (IQR)** | 1.5 (1.0, 2.0) | 1.5 (1.0, 2.0) | 0.070 | 1.5 (1.0, 2.0) | 1.5 (1.0, 2.0) | 0.070 |
| **Number of relapses in previous 2 years, mean (SD)** | 0.26 (0.56) | 0.24 (0.53) | 0.034 | 0.28 (0.57) | 0.26 (0.55) | 0.032 |
| **Duration of current treatment, years, median (IQR)** | 2.99 (2.05, 4.88) | 2.86 (2.04, 4.79) | 0.074 | 2.98 (2.03, 4.96) | 2.77 (2.00, 4.93) | 0.064 |
| **Baseline DMT efficacy group, number (%)** |  |  | 0.017 |  |  | 0.044 |
| **Platform** | 1585 (70.0%) | 198 (69.2%) |  | 1431 (71.0%) | 171 (69.0%) |  |
| **Moderate efficacy** | 679 (30.0%) | 88 (30.8%) |  | 585 (29.0%) | 77 (31.0%) |  |
| **Number of CSLs on index MRI, number (%)** |  |  | 0.278 |  |  | 0.249 |
| **1** | 1134 (50.1%) | 124 (43.4%) |  | 999 (49.6%) | 111 (44.8%) |  |
| **2** | 435 (19.2%) | 55 (19.2%) |  | 390 (19.3%) | 46 (18.5%) |  |
| **3** | 227 (10.0%) | 43 (15.0%) |  | 202 (10.0%) | 38 (15.3%) |  |
| **4 to 9** | 312 (13.8%) | 55 (19.2%) |  | 279 (13.8%) | 44 (17.7%) |  |
| **10 or more** | 156 (6.9%) | 9 (3.1%) |  | 146 (7.2%) | 9 (3.6%) |  |
| **Contrast administered for index MRI, number (%)** | 1102 (48.7%) | 124 (43.4%) | 0.373 | 1008 (50.0%) | 117 (47.2%) | 0.388 |
| **Only non-enhancing lesions, number (%)** | 677 (29.9%) | 44 (15.4%) |  | 626 (31.1%) | 41 (16.5%) |  |
| **Any contrast-enhancing lesions, number (%)** | 425 (18.8%) | 80 (28.0%) |  | 382 (18.9%) | 76 (30.6%) |  |
| **Follow-up duration, years, median (IQR)** | 4.60 (2.25, 7.06) | 4.21 (1.99, 6.35) | 0.136 | 5.09 (2.98, 7.41) | 4.81 (2.45, 6.55) | 0.151 |
| **Follow-up clinic visits per year, median (IQR)** | 2.10 (1.68, 2.79) | 2.20 (1.75, 2.89) | 0.089 | 2.17 (1.75, 2.84) | 2.22 (1.74, 2.87) | 0.054 |

All listed characteristics are at baseline except follow-up duration and follow-up clinic visits per year. Emulated trial outcome models were adjusted for baseline potential confounders. <sup>a</sup>All but 0.4% (10 of 2264 for relapse and 9 of 2016 for CDW) of participants were included in the control arm for at least the first week's emulated trial. Abbreviations: CSL, clinically silent lesion; DMT, disease modifying therapy; EDSS, expanded disability status scale; SMD, standardised mean difference, quantified using Cohen d value.

Baseline DMTs and DMT efficacy group changes during follow-up are in Supplementary Table 9. Of 286 participants who escalated DMT, 153 (53.5%) escalated to moderate-efficacy DMTs and 133 (46.5%) escalated to high-efficacy DMTs. The proportion of participants who escalated DMT within 6 months following CSLs increased from 1.9% (3 of 154) pre-2011, to 16.1% (88 of 546) between 2021-2025 (Supplementary Fig. 3). Competing outcome and censoring events, including treatment strategy deviations, are plotted in Supplementary Fig. 4. Because <20% of participants remained under follow-up beyond 4 years, follow-up in the emulated trials was censored at this timepoint. Emulated trial outcome models and metrics from censoring weight models are in Supplementary Tables 10 and 11.

The estimated 4-year cumulative incidence of relapse was lower among participants who escalated DMTs (16.8% [95% CI 12.4%-23.4%]) versus control (38.9% [95% CI 35.8%-42.1%]), risk difference (RD) −22.0% (95% CI −14.3% to −27.5%), aHR 0.34 (95% CI 0.23-0.47; Fig. 3A). Similar aHRs were estimated in participants with single (Fig. 3B) and multiple (Fig. 3C) CSLs; on both platform and moderate-efficacy baseline DMTs; and regardless of disease duration (Supplementary Fig. 5A).

**Figure 3.**
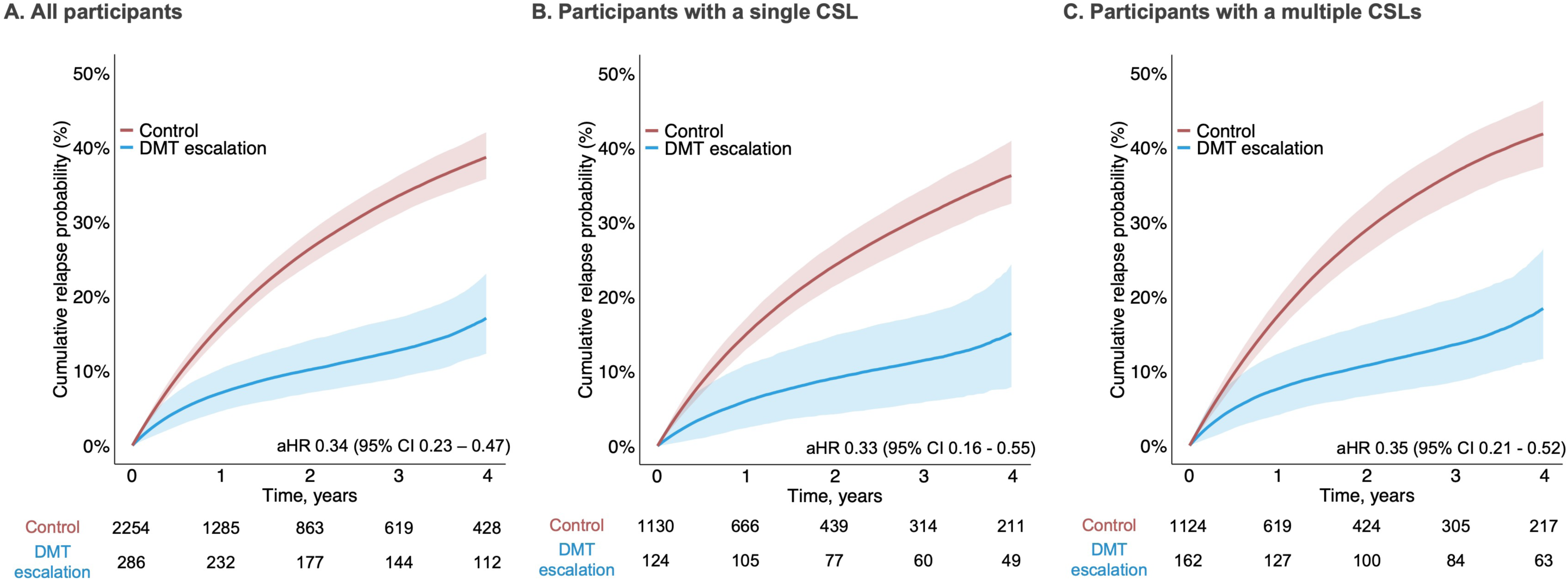
Estimated effect of DMT escalation after CSLs on the probability of relapses Cumulative probabilities (absolute risks) were generated using G-computation; adjusted hazard ratios are from pooled logistic regression models. Lines represent median probabilities, and shaded areas represent 95% confidence intervals. The control group is in red and the DMT escalation group is in blue. Plots are for (A) all participants, (B) participants with single CSLs and (C) participants with multiple CSLs. Abbreviations: aHR, adjusted hazard ratio; CSL, clinically silent lesion.

Of 2,016 participants eligible for the emulated trial assessing CDW outcomes, 248 escalated DMT within 6 months (Supplementary Fig. 2). The estimated 4-year cumulative incidence of CDW was similar in the DMT escalation (16.0% [95% CI 10.8%-22.2%]) and control (17.7% [95% CI 15.3%-20.1%]) groups, risk difference −1.7% (95% CI −7.6% to +5.1%), aHR 0.89 (95% CI 0.56-1.33) for all participants (Fig. 4A), for those with single (Fig. 4B) and multiple (Fig. 4C) CSLs, and regardless of baseline DMT efficacy (Supplementary Fig. 5B). The estimated cumulative incidence of CDW was lower following DMT escalation versus control among participants with 5–10 year disease duration, but not 0–5 or ≥10-year durations (heterogeneity p=0.001; Supplementary Fig. 5B). When CDW was subdivided into RAW and PIRA, the estimated 4-year cumulative incidence of RAW was 2.6% (95% CI 1.0%-6.4%) following DMT escalation versus 7.0% (95% CI 5.5%-8.7%) for control, aHR 0.29 (95% CI 0.11-0.75), and the estimated cumulative incidence of PIRA was 13.5% (95% CI 9.1%-20.0%) versus 10.8% (95% CI 9.2%-13.3%), aHR 1.32 (95% CI 0.82-2.14; Supplementary Fig. 6).

**Figure 4.**
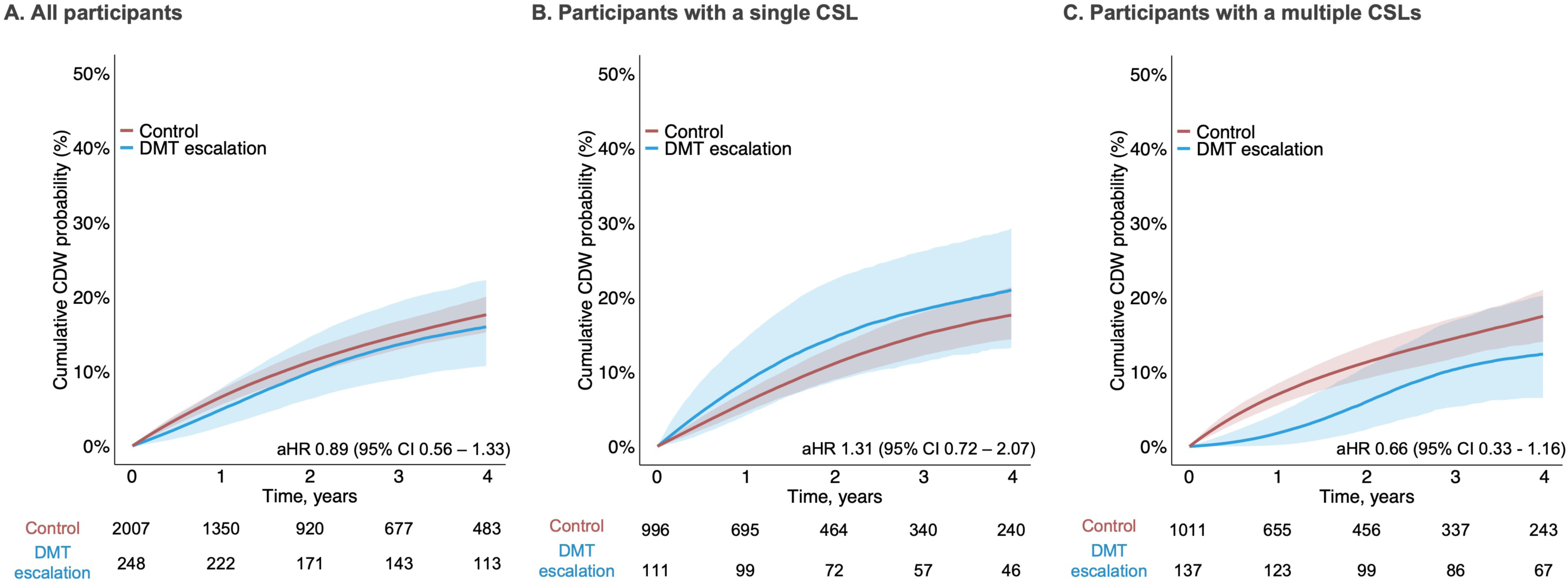
Estimated effect of DMT escalation after CSLs on the probability of confirmed disability worsening Cumulative probabilities (absolute risks) were generated using G-computation; adjusted hazard ratios are from pooled logistic regression models. Lines represent median probabilities, and shaded areas represent 95% confidence intervals. The control group is in red and the DMT escalation group is in blue. Plots are for (A) all participants, (B) participants with single CSLs and (C) participants with multiple CSLs. Abbreviations: aHR, adjusted hazard ratio; CSL, clinically silent lesion.

Compared to the main analyses, similar adjusted hazard ratios for relapse and disability worsening were estimated in sensitivity analyses: (i) adjusting models for country health expenditure; (ii) for unconfirmed disability worsening; and (iii) comparing DMT escalation to alternative control treatment strategies (Supplementary Table 12).

## Discussion

In this international registry study, adults receiving disease-modifying therapy (DMT) for RRMS who developed single or multiple clinically silent MRI lesions (CSLs) had higher subsequent 2-year risks of relapse and disability worsening than those with no CSLs. Even in recent years, most participants who developed CSLs while taking platform and moderate-efficacy DMTs did not escalate treatment. In the emulated target trial, participants who did not escalate DMTs after CSLs (unless subsequently clinically indicated) had a high estimated relapse risk: 38.9% over 4 years. DMT escalation more than halved this risk to 16.8% (aHR 0.34 [95% CI 0.23-0.47]). This effect was similar in participants with single and multiple CSLs, on platform or moderate-efficacy baseline DMTs, and irrespective of disease duration. The 4-year risk of confirmed disability worsening (CDW) was relatively low, and similar among participants who did and did not escalate DMTs.

Unlike this study, previous studies either found no association between CSLs and adverse MS outcomes,^14–16^ or only demonstrated associations if multiple (at least two^13^ or three^17–19)^ CSLs were detected. In participants with fewer CSLs, estimates of associations were often imprecise; for example, previous studies comparing participants with 1-2 CSLs to those with no CSLs reported hazard ratios for CDW of 1.33 (95% CI 0.81-2.17),^19^ and EDSS score ≥6.0 of 1.29 (95% CI 0.57-2.96).^18^ This reflects the inclusion of fewer than 2,000 participants, while this study used a large registry with >10,000 eligible participants. Previous studies (other than one^19^) examined the first 1-2 years on predominantly platform DMTs, which are infrequently initiated in contemporary practice,^6^ while participants on any licensed DMT for any treatment duration were eligible for this study. Furthermore, unlike most previous studies,^13–17,19^ a “re-baseline scan” ensured that the CSL group was not diluted by lesions emerging during DMTs’ therapeutic lag periods.^7,26^

The proportion of participants who escalated treatment in the 6 months following even contemporary CSLs was low – 16.1% since 2021 – mirroring the low proportion in a recent European cohort study (22.1% at 12 months)^13^; this may reflect a lack of randomised trial evidence. An observational study found associations between escalating DMTs and improved relapse and disability outcomes only following multiple CSLs.^13^ However, only three participants escalated DMT after a single CSL and the study was not designed to infer causation. Although our study was not randomised, several factors support a causal interpretation of the reported effects of DMT escalation. We addressed a clear clinical question by comparing explicitly defined treatment strategies following one or more CSLs. Our eligibility criteria ensured that included participants were treated in centres with access to higher-efficacy DMTs at the time of inclusion. Since implementing DMT escalation often takes several months, sequential emulation was used to temporally align eligibility, treatment allocation, and the start of follow-up, eliminating the risk of bias due to immortal time.^24,32^ Follow-up of control participants was censored if they escalated DMT without a clinical event, or did not escalate DMT within 6 months of a clinical event, with adjustment for this censoring using inverse-probability weighting based on time-updated covariates. This enabled clear separation of the studied treatment strategies and ensured that the control group did not act as a proxy for a conservative treatment approach to clinical events.

Although the 4-year risk of CDW was similar in both treatment groups, when CDW was subdivided, the risk of relapse-associated worsening (RAW) was lower with versus without DMT escalation, while the risk of progression independent of relapse activity (PIRA) was somewhat higher, although the between-group difference was imprecisely estimated. These results reflect the mechanism of current DMTs, which ameliorate acute CNS inflammation but not other drivers of disability progression.^27,33^ Treatment effects on overall CDW may not be detected over relatively short follow-up, given the low rate of CDW in contemporary treated RRMS cohorts.^28^ Nevertheless, the absence of a discernible effect of DMT escalation following CSLs on subsequent disability worsening exposes a potential limitation of treatment escalation strategies (with initial use of platform or moderate-efficacy DMTs) for RRMS. The alternative strategy of early high-efficacy DMT use was associated with favourable disability outcomes in previous observational studies^2,4,5^ and is currently being tested against treatment escalation approaches in randomised trials.^34,35^

### Limitations

This study has several limitations. First, it did not investigate adverse effects of escalating DMTs, since safety data in MSBase is currently sparse, so the findings must be interpreted in the context of the known risks of higher-efficacy DMTs. Second, while confounding was addressed by adjusting for many important prognostic factors that drive treatment decisions, unmeasured confounding cannot be excluded. The decision to escalate treatment may have occurred more frequently in participants with unrecorded predictors of disability progression, such as infratentorial or spinal lesions,^19^ diminishing the estimated effect of DMT escalation on disability worsening. Third, a real-world registry with non-standardised MRI lesion counts was used. This reflects current clinical practice, but if artificial intelligence tools are adopted to improve CSL detection sensitivity,^36^ further studies will be required to understand the clinical implications of the additional CSLs that are detected. Fourth, missing MRI data reduced the number of eligible participants. However, following primary source review by local investigators, participants from 99 clinics in 26 countries were included, suggesting that the results have broad generalisability. Fifth, spinal MRIs were not studied; however spinal CSLs are rarely identified in the absence of concurrent brain CSLs,^37^ and current international guidelines do not recommend routine spinal MRI monitoring.^7^ Sixth, insufficient data were available to stratify analyses by CSL anatomical location. Infratentorial lesions are a poor prognostic marker,^19^ but whether their management should differ from supratentorial lesions remains unstudied. Finally, a higher proportion of participants with versus without CSLs escalated DMTs during follow-up, which may have attenuated associations between CSLs and outcomes (assessed at 2 years), and precluded meaningful assessment of longer-term outcomes. The impact of DMT escalation was assessed over 4 years’ follow-up, since beyond that time <20% of participants remained under follow-up.

## Conclusion

People with RRMS who develop single or multiple clinically silent lesions on treatment-monitoring MRI brain scans have higher subsequent risks of relapses and disability worsening than people without CSLs. DMT escalation after single or multiple CSLs more than halves the 4-year relapse risk, but disability worsening occurs at a similar rate regardless of DMT escalation. These findings oppose the concept of a tolerable radiological “minimal evidence of disease activity”.^17,18,20^ Hence, contrary to clinical guidelines,^8–10^ and alongside careful consideration of these therapies’ risks, detection of even a single CSL should prompt consideration of DMT escalation.

## Data availability

Deidentified data from the participating cohort and data dictionary can be requested from the principal investigators, conditional after obtaining approvals from the appropriate institutional review boards. The MSBase registry is a data processor and warehouses data from individual principal investigators who agree to share their datasets on a project-by-project basis. Data access to external parties can be granted on reasonable request at the sole discretion of the principal investigators, who will need to be approached individually for permission.

## Author contributions

Cyrus Daruwalla and William Brown had full access to all the data in the study and take responsibility for the integrity of the data and the accuracy of the data analysis.

William Brown, Jonathan Sterne, and Tomas Kalincik contributed equally and are joint last authors.

*Concept and design:* Daruwalla, Brown, Sterne, Kalincik, Coles, and Madley-Dowd.

*Acquisition, analysis, or interpretation of the data:* All authors.

*Drafting of the manuscript:* Daruwalla, Brown, Sterne, Kalincik, Coles and Madley-Dowd.

*Critical revision of the manuscript for important intellectual content:* All authors.

*Statistical analysis:* Daruwalla, Brown, Sterne, Kalincik, Coles, Madley-Dowd, Roos, and Kremler.

*Obtained funding*: Daruwalla, Brown, Kalincik and Coles.

*Administrative technical or material support*: Daruwalla, Brown, Sterne, Kalincik, Coles, Madley-Dowd, Roos, Cunniffe, Kremler, Tolkovsky, and Wardle.

*Supervision*: Brown, Sterne, Kalincik, Coles, Madley-Dowd, Roos, and Cunniffe.

## MSBase co-investigators and contributors

The following contributors participated in data acquisition: From Neuroimmunology Centre, Department of Neurology, Royal Melbourne Hospital, Melbourne, Australia, Dr Mark Marriott, Dr Trevor Kilpatrick, Dr John King, Dr Katherine Buzzard, Dr Ai-Lan Nguyen, Dr Chris Dwyer, Dr Vivien Li, Dr Prashanth Ramachandran, Dr Melissa Chu, Ms Lisa Taylor, Ms Katherine Fazzolari, Ms Hasini Fernando, Ms Margaret Bolger, Ms Jersey Higgins. From Department of Medical and Surgical Sciences and Advanced Technologies, GF Ingrassia, Catania, Italy, Dr Clara Chisari, Dr Lo Fermo Salvatore. From CHUM and Universite de Montreal, Montreal, Canada, Dr Catherine Larochelle. From Academic MS Center Zuyd, Department of Neurology, Zuyderland Medical Center, Sittard-Geleen, Netherlands, Dr Raymond Hupperts. From Centro de Esclerosis Múltiple, Hospital Universitario Nacional de Colombia, Colombia, Juan Camilo Carrillo. From Hospital Universitario Donostia and IIS Biodonostia, San Sebastián, Spain, Dr Javier Olascoaga. From Department of Neurology, Second Faculty of Medicine, Charles University and Motol University Hospital, Prague, Czech Republic, Jana Libertinova. From Department of Neurology, Faculty of Medicine in Pilsen, Charles University and University Hospital Pilsen, Pilsen, Czech Republic, Marek Paterka. From Department of Neurology and Centre of Clinical Neuroscience, First Faculty of Medicine, Charles University and General University Hospital in Prague, Prague, Czech Republic, Marta Vachova. From Neuroimmunology Centre, Department of Neurology, Neuroimmunology Centre, Department of Neurology, Royal Melbourne Hospital, Melbourne, Australia, Tomas Kalincik. From Neurology Institute, Harley Street Medical Center, Abu Dhabi-UAE, Bassem Yamout. From Service de Neurologie, CISSS Chaudière-Appalache, Levis, Canada, Pierre Grammond. From Department of Neurology, Hospital Universitario Virgen Macarena, Sevilla, Spain, Sara Eichau. From Hospital Universitario Virgen Macarena, Sevilla, Spain, Guillermo Izquierdo. From Azienda Ospedaliera di Rilievo Nazionale San Giuseppe Moscati Avellino, Avellino, Italy, Daniele Spitaleri. From St. Michael’s Hospital, Toronto, Canada, Jiwon Oh. From Department of Neurology, University Hospital Ostrava, Ostrava, Czech Republic, Pavel Hradilek . From First Department of Neurology, Masaryk University, St. Anne’s University Hospital, Brno, Czech Republic, Michal Dufek . From Neurology Department, Wexham Park Hospital, Frimley Health Foundation Trust, Slough, United Kingdom, Silvia Messina. From Neurology Unit, AST Macerata, Macerata, Italy, Elisabetta Cartechini. From St Vincent’s Hospital, Sydney, Australia, Jennifer Massey. From College of Medicine and Public Health, Flinders University, Adelaide, Australia, Mark Slee. From Department of Neurology, Unidade Local de Saúde de São João,Porto, Portugal, Joana Guimarães. From Centro Hospitalar Universitario de Sao Joao, Porto, Portugal, Maria Edite Rio. From Neurology Unit, Hospital General Universitario de Alicante, Alicante, Spain, Angel Perez sempere. From Neurology Institute and MS Center, Harley Street Medical Centre, Abu Dhabi, United Arab Emirates, Bassem Yamout. From Department for Neurology, Department of Neurology, University Medical Centre Ljubljana, Slovenia, Gregor Brecl Jakob. From AZ Alma Ziekenhuis, Sijsele - Damme, Belgium, Danny Decoo. From Centro Internacional de Restauracion Neurologica, Havana, Cuba, Jose Antonio Cabrera-Gomez. From Department of Neurology, Concord Repatriation General Hospital, Sydney, Australia, Todd A. Hardy. From Neurosciences Department, Mater Dei Hospital, Birkirkara, Malta, Marija Cauchi. From Neurology, Az Sint-Jan Brugge, Bruges, Belgium, Melissa Cambron. From Department of Neurology, McGill University, Montreal, Canada, Fraser Moore. From Hospital Universitario de la Ribera, Alzira, Spain, Jose Andres Dominguez. From Department of Neurology, , AHEPA University Hospital, Thessaloniki, Greece, Nikolaos Grigoriadis. From Department of Neurology, Graduate School of Medical Sciences, Kyushu University, Fukuoka City, Japan, Noriko Isobe. From Department of Neurology, 3rd Faculty of Medicine, Charles University and University Thomayer Hospital, Prague, Czech Republic, Jan Mares. From Department of Neurology, Tokyo Metropolitan Health and Medical Treatment Corporation Ebara Hospital, Department of Neurology, Tokyo, Japan, Chiyoko Nohara. From Neurology, Nagoya City University School of Medical Sciences, Neurology, Aichi Prefecture, Japan, Masayuki Mizuno. From Department of Neurology and Clinical Neuroscience, Yamaguchi University Graduate School of Medicine, Department of Neurology and Clinical Neuroscience, Ube, Japan, Fumitaka Shimizu. From Neurology and neurosurgery, Riga East Clinical University Hospital, Neurology and neurosurgery, Riga, Latvia, Guntis Karelis. From Department of Neurology, School of Medicine and Koc University Research Center for Translational Medicine (KUTTAM), Koc University, School of Medicine, Istanbul, Turkey, Ayse Altintas. From Multiple Sclerosis Research Center, Neuroscience Institute, Multiple Sclerosis Research Center, Neuroscience Institute, Tehran University of Medical Sciences, Tehran, Iran, Abdorreza Naser Moghadasi. From Neurology Department, King Fahad Specialist Hospital-Dammam, Saudi Arabia, Talal Al-Harbi. From Department of Neuroscience, Hospital Germans Trias i Pujol, Badalona, Spain, Cristina Ramo-Tello. From Veszprém Megyei Csolnoky Ferenc Kórház zrt., Veszprem, Hungary, Imre Piroska. From Centro de Esclerosis Múltiple de Buenos Aires (CEMBA), Buenos Aires, Argentina, Edgardo Cristiano. From Neuroscience Department, Barwon Health, University Hospital Geelong, Geelong, Australia, Cameron Shaw. From St Vincents Hospital, Fitzroy, Melbourne, Australia, Neil Shuey. From Townsville Hospital, Townsville, Australia, Mike Boggild. From Department of Neurology, CSSS Saint-Jérôme, Saint-Jerome, Canada, Julie Prevost. From Neurology department, Booalisina Hospital, Mazandaran University of Medical Sciences, Sari, Iran, Seyed Mohammad Baghbanian. From Neurology, St. Marianna Univertisy School of Medicine, Sugao, Miyamae, Kawasaki, Kanagawa, Japan , Kenzo Sakurai. From Hospital Angeles de las Lomas. Instituto Mexicano de Neurociencias., Huixquilucan Estado de Mexico, Mexico, Eli Skromne. From Department of Neurology, Christchurch Hospital, Christchurch, New Zealand, Deborah Mason. From Department of Neurology, Royal Hospital, Muscat, Oman, Jabir Alkhaboori. From University of Medicine and Pharmacy Victor Babes Timisoara, Mihaela Simu. From Dept.of Neurological Sciences, Faculty of Medicine, Yeditepe University, Istanbul, Turkey, MSBase Centre Custodian. From Department of Neurology, Haydarpasa Numune Training and Research Hospital, Istanbul, Turkey, Recai Turkoglu. From Royal North Shore Hospital, Sydney, Australia, John Parratt. From Neuro Clinic, Neurology, Cairo, Egypt, Nevin Shalaby. From Department of Neurology, Semmelweis University, Budapest, Hungary, Magdolna Simo. From Péterfy Sandor Hospital, Budapest, Hungary, Krisztina Kovacs. From Department of Neurology and Stroke, BAZ County Hospital, Miskolc, Hungary, Attila Sas. From Neurology Department, Szent Borbála Kórház, Tatabánya, Hungary, Andrea Mike. From Department of Neurology, Brain and Nerve Center, Fukuoka Central Hospital, Department of Neurology, Brain and Nerve Center, Fukuoka, Japan, Yuri Nakamura . From Department of Neurology, Waikato Hospital, Hamilton, New Zealand, Beatriz Romero Ferrando. From Neurology, Ankara University Ibni Sina Hospital, Canan Yucesan. From Neurology, Kyiv Medical University, Ukraine, Oksana Kopchak.

## Other contributors

Administrative and technical support was provided by: From the Cambridge Clinical MS Research Group Ms Toni Bray, Ms Jacky Porter, Mr Nosson Almeida, Ms Magdalena Dragan, Ms Nadira Fernando, Ms Samantha Grant, Mr Keith Logan, Ms Debbie Morrall and Ms Maria Sanfilippo; and from the MSBase Operations team Ms Charlotte Sartori, Ms Pamela Farr, Ms Rein Moore, Mr Dusko Stupar, Ms Cynthia Tang, Ms Sonya Smirnova, and Ms Abeer Khimani.

