## Supplementary material for "Treatment escalation after clinically silent MRI lesions in relapsing-remitting multiple sclerosis"

### Supplementary Methods

#### Data quality checking procedure

- Duplicate patient records were removed.
- Centres with <10 patient records were excluded.
- Patients with missing date of birth were excluded.
- MS onset dates after the data extract date were removed.
- Patients with missing date of the first clinical presentation of MS were excluded.
- The dates of MS onset and the first recorded MS course were aligned.
- Patients with the age at onset outside the 0–100 range were excluded.
- A logical sequence of the MS courses (i.e. clinically isolated syndrome, then relapsing-remitting MS, then secondary progressive MS) was assured.
- Visits with a missing visit date or the recorded date before the clinical MS onset or after the date of data extract were removed.
- EDSS scores outside the range of possible EDSS values were removed.
- Duplicate visits were merged.
- MS relapses with a missing onset date or the recorded date after the date of data extract were removed.
- Duplicate MS relapses were merged.
- Relapses occurring within 30 days of each other were merged.
- Therapies with erroneous date entries were removed (e.g. commencement date after the termination date, commencement after the data extract date, commencement of disease modifying therapy before the year 1980).
- Duplicate treatment entries were removed.
- Where multiple disease modifying therapies were recorded with overlapping dates, the treatment end date of the earlier therapy was imputed as the commencement date of the following therapy.
- For induction therapies (alemtuzumab, cladribine, and autologous haematopoietic stem cell transplant), treatment end date was imputed as the day before commencement of the subsequent therapy, if present.

#### Eligibility requirements

- Diagnosed with relapsing-remitting multiple sclerosis (RRMS) according to the applicable diagnostic criteria at the time of diagnosis.
- Onset of symptoms aged 16 years and over.
- Presence of the complete MSBase minimum dataset (sex, date of birth, date of clinical onset, and dates of relapses).
- At least one consecutive pair of MRI head scans between 6 and 18 months apart, henceforth referred to as the "comparator MRI" and subsequent "index MRI", where at least one index MRI fulfilled all of the following requirements:
    - Performed on or after 1st January 2007 (so that the study reflects the modern treatment era, including the availability of highly effective DMTs; natalizumab was the first such DMT to receive a European Medicines Agency marketing authorisation in 2006[2]) and before 1st October 2024 (6 months prior to the date of data extraction, to ensure that the post-MRI treatment strategy was discernible for all included participants).
    - Performed after the availability of a DMT of higher efficacy than the participant's baseline DMT in their treating centre, the date of which was determined as the first date of initiation of such a DMT for any participant treated at their centre, as recorded in MSBase. This ensured that the positivity assumption was satisfied.
    - Performed before the diagnosis of secondary progressive MS.
    - So that the presence or absence and number of clinically silent lesions was discernible, had recorded data on at least one of: (a) the number of new or enlarging T2-hyperintense lesions on the index MRI; (b) the total number of T2-hyperintense lesions on both the index MRI and the comparator MRI; or (c) ≥1 T1 gadolinium contrast-enhancing lesions on the index MRI.
    - On a single DMT throughout the interscan interval with the first dose given at least 6 months prior to the comparator MRI for all DMTs except cladribine and alemtuzumab, and at least 18 months prior to the comparator MRI for cladribine and alemtuzumab (since these DMTs require two courses separated by 12 [alemtuzumab] or 13 months [cladribine]).
    - Presence of a recorded EDSS score within 12 months prior to the comparator MRI scan, henceforth referred to as the "baseline EDSS score".

- Performed in clinically silent periods, i.e. no relapses or EDSS scores that were higher than the baseline EDSS score recorded between the date of the baseline EDSS score and one week after the index MRI; this additional week was required to enable allocation to the studied treatment strategies for the first of the weekly emulated trials for all participants, prior to them having the outcome of interest.
    - At least one follow-up clinic visit after the index MRI scan. Additionally, for analyses of 6-month confirmed disability worsening outcomes, at least two recorded post-baseline EDSS scores at least 6 months apart.
    - Not enrolled in a randomised controlled trial at the time of the index MRI scan, or during the follow-up period.
- At least one eligible participant in their country of care had the outcome of interest during follow-up.

Included index MRI scans were determined to have clinically silent lesions (CSLs) if one or more of the following applied: (a) ≥1 new or enlarging T2-hyperintense lesions were recorded; (b) the total number of T2-hyperintense lesions exceeded that of the comparator MRI scan; or (c) ≥1 contrast-enhancing lesion was recorded.

For each eligible participant, their earliest eligible scan pair was included for the comparison of prognostic outcomes in participants with and without CSLs, and their earliest scan pair demonstrating CSLs while taking a platform or moderate-efficacy DMT was included for the emulated trial of DMT escalation.

**DMT escalation definition**

DMT escalation was defined as a change from either:

a) Platform DMT to moderate-efficacy DMT, or
b) Platform DMT to high-efficacy DMT, or
c) Moderate-efficacy DMT to high-efficacy DMT

**Outcome definitions**

Relapse: the occurrence of new or worsened symptoms typical of an acute central nervous system demyelinating event, with a duration of at least 24 hours, in the absence of fever or infection, and occurring at least 30 days after the previous relapse.[1]

6 month confirmed disability worsening (CDW): an increase in EDSS score of at least 1.5 steps if the baseline EDSS score was 0; at least 1 step if the baseline EDSS score was between 1.0 and 5.5, inclusive; or at least 0.5 steps if the baseline EDSS score was ≥6.0, confirmed at another assessment at least 6 months later (as well as any assessments within 6 months after the increased EDSS score, if performed), with the confirmatory EDSS score not within 30 days after a relapse.[2] The date of the initial EDSS score increase is used as the date of CDW.

Progression independent of relapse activity (PIRA): A 6 month CDW event, defined as above, in the absence of any relapses between the baseline EDSS score (defined as per the study baseline EDSS score, and re-baselined if there was a later EDSS improvement to the new EDSS score, or if there was a later relapse, re-baselined to the first recorded EDSS score at least 30 days after the relapse) and the CDW event.[3]

Relapse-associated worsening (RAW): A 6 month CDW event, defined as above, in the presence of a relapse between the baseline EDSS score and the CDW event. This corresponded to any CDW event that did not satisfy the definition of PIRA.

CDW and PIRA outcomes were identified using the MSoutcomes R package (version 0.2.1).[4]

**Statistical analysis**

Prognosis following clinically silent lesions

The cumulative incidence of outcomes was derived from unadjusted Kaplan-Meier estimates. Cox proportional hazards models were used to compare hazards of outcomes in participants with or without CSLs. These models were stratified by country of care (grouping countries with <40 participants) and adjusted for the following potential baseline confounders, determined at the time of the index MRI scan:

- Sex
- Age (centred on the median age)

- Disease duration
- Calendar year (centred on the earliest year [2007])
- EDSS score
- Number of relapses in the prior 2 years
- DMT efficacy group

Where there was a non-linear relationship between a continuous covariate and the risk of relapse, the lowest order polynomial term with a good fit to the data was used (as determined by visual inspection of plots of each covariate against the empirical relapse risk, overlayed with the modelled linear, quadratic and cubic risks) in all models (including Cox models for prognosis analyses, and outcome and weighting models for emulated trials): quadratic terms for age, disease duration and calendar year and cubic terms for EDSS score.

*Alternative comparisons and subgroup analyses:*
As well as comparing all participants with and without CSLs, participants without CSLs were compared to participants with 1 or ≥2 CSLs, and with contrast-enhancing and non-enhancing CSLs (restricted to participants who received gadolinium contrast on their index MRI scan). Subgroup analyses explored variation in associations with the baseline DMT efficacy group (platform, moderate-efficacy or high-efficacy), and baseline disease duration (0-5 years, 5-10 years, or ≥10 years). Heterogeneity between subgroup log hazard ratios was tested using Cochran's Q test.

*Sensitivity analyses:*
First, a sensitivity analysis was performed replacing country stratification with adjustment of models for health expenditure in the country of care (categorised according to World Health Organisation global health expenditure data, 2016[5] as <1500, 1500-3000 or ≥3000 US dollars per capita per annum). Second, a sensitivity analysis was conducted in the subset of participants who had a total (old and new) T2 lesion count recorded on their index MRI scan, with risks of outcomes analysed both with and without the total lesion count as a covariate in the models; this aimed to test whether the presence of new CSLs predicted outcomes over and above the total burden of T2 lesions.[6] Third, unconfirmed disability worsening outcomes were analysed, requiring the same EDSS score increase as the CDW definition, but without a requirement for later confirmation of the increased EDSS score.

<u>Sequentially emulated trial of DMT escalation following CSLs</u>

The target trial specification and emulation are outlined in Supplementary Table 1.

*Treatment strategies*
Starting each week for 6 months following the index MRI, the following non-blinded treatment strategies were compared:

1. *DMT escalation*
2. *Control*: no DMT escalation unless a clinical event (relapse for CDW outcome analyses, disability worsening [an EDSS score increase defined as per CDW, but without the requirement for later confirmation] for relapse outcome analyses) occurs, in which case DMT escalation must occur during the next 6 months.

The control strategy was designed to ensure that this group did not act as a proxy for a conservative approach to clinical events that occur during follow-up, by requiring DMT escalation in response to such events. For this purpose, for relapse outcome analyses, clinical events were defined as disability worsening, but not relapse (since the time-to-event nature of the outcome meant that participants would be censored when they had a relapse) and vice versa.

*Sequential trial emulation protocol*

1. Identify censoring and outcome events:
    - For participants in the control arm in at least one baseline week (i.e. all participants except those who escalate DMT in the first week after the CSL), identify the earliest of the following competing events:
        - The outcome
        - DMT escalation despite no clinical event (disability worsening for relapse outcomes, relapse for CDW outcomes) in the preceding 6 months
        - 6 months after a clinical event with no subsequent DMT escalation
        - The last clinic visit

   - For participants in the DMT escalation arm in at least one baseline week (i.e. participants who escalate DMT in the first week after the CSL, or participants initially in the control arm whose first competing event is DMT escalation despite no clinical event and this occurs within 6 months after the CSL), identify the earliest of the following competing events:
     - The outcome
     - The last clinic visit
2. Transform data on included participants into person-week format:
   - Generate new row for each week that the participant is eligible to remain included.
   - Generate a column with the time-updating treatment arm during each week. Participants who escalate DMT in the first week are in the DMT escalation arm from the first week. All other participants remain in the control arm until the week that they are eligible to enter the DMT escalation arm, defined as above.
   - Generate columns with time-updating data on each potential confounding variable, using the most recent recorded entry of each covariate on or prior to the given week.
   - Generate a column with a time-updating eligibility indicator, identifying weeks in which a participant is eligible to enter a new baseline week. Participants who enter the DMT escalation arm in any baseline week are no longer eligible for either arm in subsequent weeks.
3. Transform the data into trial-week format:
   - Expand the person-week data so that for each week in which a participant is eligible to enter a "trial" baseline week, the row corresponding to that and all subsequent weeks is replicated by the number of times that week will be included. Each person-trial is given a unique ID.
   - For each person-trial, rows corresponding to weeks before trial entry are removed, and a trial-week variable corresponding to the number of weeks after entry into the trial is generated.
   - For each person-trial, rows following censoring are removed, as determined in step 1.
   - Baseline covariates for each trial correspond to the time-varying covariate recorded in the baseline week of that trial.
4. For participants in the control arm, calculate inverse probability of censoring weights (IPCW) for censoring due to DMT escalation despite no clinical event (IPCW1), as follows:
   - For participants who are in the control arm for at least one baseline week, restrict the person-week data to weeks in which this reason for deviation from the baseline treatment strategy is possible (i.e. remove weeks in the 6 months after a clinical event, and any weeks after censoring or DMT escalation).
   - Label weeks in which participants are censored due to DMT escalation despite no clinical event in the preceding 6 months.
   - Calculate the denominator of IPCW1 for each week using logistic regression to model the probability of remaining free from this reason for censoring in each week post MRI, based on the following time-updating covariates:
     - Sex
     - Age (centred on the median age; linear and quadratic terms included)
     - Disease duration (linear and quadratic terms included)
     - Calendar year (centred on the earliest year [2007]; linear and quadratic terms included)
     - EDSS score (linear, quadratic and cubic terms included)
     - Number of relapses in the prior 2 years
     - Country of care (grouping countries with <40 participants)
     - DMT efficacy group
     - Weeks post MRI
     - Number of CSLs on the index MRI scan (categorised as 1, 2, 3, 4-9 or ≥9)
     - Contrast enhancing lesions on the index MRI scan (categorised as present, absent, or contrast not given)
     - Follow-up MRI brain scan results in the previous 6 months (categorised as the presence or absence of new/enlarging/enhancing lesions, or no scan performed)
   - Weeks in which censoring for this reason is impossible are assigned an IPCW denominator of 1.
   - Calculate the numerator used to stabilise IPCW1 using another logistic regression model which models the probability of remaining free from this reason for censoring in each week post MRI based on just the week post MRI and calendar year.
   - For each week of each person-trial, IPCW1 is the cumulative product of the IPCW1 numerator divided by the IPCW denominator up to that person-week.

5. For participants in the control arm, calculate inverse probability of censoring weights for censoring due to no DMT escalation despite a clinical event 6 months earlier (IPCW2), as follows:
    - For participants who are in the control arm for at least one trial, restrict the person-week data frame to weeks in which this reason for deviation from the baseline treatment strategy is possible (i.e. only weeks in the 6 months after a clinical event).
    - Label weeks in which participants are censored due to no DMT escalation despite a clinical event in 6 months earlier.
    - The remaining steps are identical to those for IPCW1, from calculating the IPCW denominator onwards.
6. Calculate the final IPCW for each week of each person-trial as the product of IPCW1 and IPCW2.
7. Model the outcome to generate risks and hazard ratios
    - Fit a logistic regression model of the outcome on the trial-week data, weighted using the final IPCW, and adjusted for the value of each of the baseline week covariates (as per covariates in the models for the IPCW denominators listed above).
8. Use bootstrapping with 1000 samples to generate confidence intervals
    - Repeat steps 1-7 above 1000 times, each time using datasets generated by randomly sampling the included participants with replacement.
    - The 2.5$^{th}$ and 97.5$^{th}$ percentile of the hazard ratios generated by these bootstrapped analyses are used as the 95% confidence interval.

*G computation protocol to generate absolute risks and cumulative incidence curves:*

1. Duplicate the trial-week data and artificially expand data for each participant-trial to include rows for weeks after censoring up to the maximum follow-up duration (208 weeks).
2. Artificially assign every participant to the DMT escalation arm in one copy of the data, and the control arm in the other copy.
3. Fit the logistic regression outcome model with the addition of product terms between the treatment arm and the number of weeks post baseline (both as linear and quadratic terms) to allow the effect of treatment to vary over time.
4. Use the predicted values from these models to estimate 4-year absolute risks and plot estimates of the cumulative incidence of outcomes over time had every participant followed each of the treatment strategies.

*Subgroup analyses*

Analyses were performed in subgroups based on the number of CSLs (1 or ≥2), baseline DMT efficacy groups (platform or moderate-efficacy) and disease duration (0-5 years, 5-10 years, or ≥10 years). Outcome and censoring weight models for subgroup analyses were adjusted for health expenditure in the country of care (as in the first sensitivity analysis, detailed below) instead of a marker of the country of care, since subgroups did not all include participants from each country with the outcomes of interest. We tested for heterogeneity between subgroup log hazard ratios using Cochran's Q test. Subgroups based on contrast enhancement of CSLs (contrast-enhancing or non-contrast-enhancing lesions) were attempted, but confidence intervals were asymmetrical and likely to be unreliable due to failure of model convergence in a proportion of bootstrap samples; these results are therefore not presented.

*Sensitivity analysis*

First, a sensitivity analysis was performed replacing adjustment for the country of care with adjustment of models for health expenditure in the country of care (categorised according to World Health Organisation global health expenditure data, 2016[5] as <1500, 1500-3000 or ≥3000 US dollars per capita per annum). Second, unconfirmed disability worsening outcomes were analysed, requiring the same EDSS score increase as the CDW definition, but without a requirement for later confirmation of the increased EDSS score. Third, DMT escalation was compared to alternative control treatment strategies: (a) without requiring DMT escalation following clinical events (hence without IPCW1); and (b) additionally without requiring unchanged DMT if there is no clinical event (hence without IPCW1 and IPCW2; analogous to the intention-to-treat effect).

### Supplementary tables

**Supplementary Table 1. Specification of the target trial of DMT escalation following clinically silent lesions and its emulation**

| Protocol Component | Target trial specification | Target trial emulation |
|---|---|---|
| **Eligibility criteria** | • Diagnosed with relapsing-remitting multiple sclerosis (RRMS)<br>• Aged 16 years or older at the time of the first MS symptom<br>• Had an MRI head scan within the last 6 months (referred to as the index MRI scan) demonstrating ≥1 CSL when compared to an MRI head scan between 6 and 18 months previously (referred to as the comparator MRI scan).<br>• Taking any platform or moderate-efficacy DMT continuously, starting at least 6 months (or 18 months for cladribine and alemtuzumab) prior to the comparator MRI and continuing at least until the index MRI<br>• No relapses or EDSS score increases between the comparator MRI and 1 week after the index MRI | • Same as target trial with the following modifications:<br>• The index MRI must have occurred after 31/12/2006, at least 6 months prior to the date of data extraction, and after the availability of a DMT of higher efficacy than the participant's baseline DMT in their treating centre. |
| **Treatment strategies** | a. Escalate DMT<br>b. Control: Do not escalate DMT unless there is a clinical event (relapse or disability worsening), in which case the DMT must be escalated within 6 months of this clinical event. | Same as for the target trial |
| **Assignment procedures** | Random assignment to a treatment strategy.<br>Participants and clinicians are not blinded to the treatment strategy. | Non-random assignment to a treatment strategy. A sequence of target trials starts each week for 6 months after the index MRI. In each week, the DMT escalation arm comprised participants who escalated DMT in the prior week and had not had a relapse or disability worsening event since the index MRI; the control arm comprised all other remaining participants.<br>To balance characteristics between treatment arms, outcomes are adjusted for the following baseline covariates:<br>• Sex<br>• Age<br>• Disease duration (from first MS symptom)<br>• EDSS score<br>• Number of relapses in the previous 2 years |

| **Protocol Component** | **Target trial specification** | **Target trial emulation** |
|---|---|---|
| | | • DMT efficacy group<br>• Country of care (grouping countries with <40 participants)<br>• Number of CSLs on the index MRI scan (categorised as 1, 2, 3, 4-9 or ≥9)<br>• Contrast enhancing lesions on the index MRI scan (categorised as present, absent, or contrast not given)<br>• Calendar year<br>• Number of weeks after the index MRI<br>Participants and clinicians are not blinded to the treatment strategy. |
| **Follow-up** | For each eligible individual, follow-up begins at the time of treatment assignment and continues until the earliest of development of the outcome of interest, loss to follow-up, or 4 years after the index MRI scan. | For each person-trial, follow-up begins at the time of treatment assignment and continues until the earliest of development of the outcome of interest, deviation from the baseline treatment strategy, the date of the last clinic visit, or 4 years after the index MRI scan. |
| **Outcomes** | Primary: Relapse<br>Secondary: 6 month confirmed disability worsening | Same as for the target trial |
| **Causal contrasts** | Intention-to-treat effect<br>Per-protocol effect<br><br>Effect measures: 4-year absolute risks and hazard ratios. | Observational analogue of the per-protocol effect. Participants in the control arm are censored if they deviate from their assigned treatment strategy. Outcomes are weighted on cumulative inverse probabilities of remaining uncensored, modelled using logistic regression models accounting for time-updated covariates (mirroring the baseline covariates listed above [in the Assignment procedures row], with the addition of a time-updating MRI covariate [categorised according to whether an MRI scan in the prior 6 months showed new lesions, did not show new lesions, or was not performed]).<br><br>Effect measures: 4-year absolute risks (estimated using G-computation) and hazard ratios. |
| **Identifying assumptions** | Validity of the estimates assumes that selection bias is not introduced by time-varying confounding due to between-arm discrepancies in loss-to-follow-up and, for the per-protocol effect, deviation from the assigned treatment strategy. | The per-protocol effect is valid assuming (a) no unmeasured baseline confounding and (b) no unmeasured time-varying confounding introduced by artificial censoring on deviation from the control treatment strategy or loss to follow-up. |

**Supplementary Table 2. Number of included patients per centre**

| Centre | Country | Patients (n) |
|---|---|---|
| Azienda Ospedaliero Universitaria Policlinico G. Rodolico - San Marco | Italy | 790 |
| University hospital Motol Prague | Czechia | 651 |
| Izmir University of Economics Medical Point Hospital | Turkey | 606 |
| Fakultni nemocnice Plzen | Czechia | 480 |
| KTU Medical Faculty Farabi Hospital | Turkey | 469 |
| Hospital Teplice | Czechia | 466 |
| John Hunter Hospital | Australia | 398 |
| The Royal Melbourne Hospital | Australia | 391 |
| S. Maria delle Croci Hospital of Ravenna | Italy | 352 |
| American University of Beirut Medical Center | Lebanon | 351 |
| Alfred Health | Australia | 319 |
| Fakultni nemocnice Olomouc | Czechia | 304 |
| University Hospital Center Zagreb | Croatia | 246 |
| Hospital Virgen De La Arrixaca | Spain | 230 |
| Amiri Hospital | Kuwait | 223 |
| Chaudière-Appalaches Integrated Health and Social Services Center | Canada | 197 |
| Hospital Clinic de Barcelona | Spain | 185 |
| University Hospital Ghent | Belgium | 178 |
| Box Hill Hospital | Australia | 177 |
| South East Trust | UK | 176 |
| Hospital Universitario Virgen Macarena | Spain | 169 |
| AORN San Giuseppe Moscati Avellino | Italy | 166 |
| St Andrews Place | Australia | 162 |
| Garibaldi Hospital | Italy | 157 |
| CHUM - Hopital Notre Dame | Canada | 150 |
| Addenbrooke's Hospital | UK | 141 |
| Hospital de Galdakao-Usansolo | Spain | 125 |
| Cliniques Universitaires Saint-Luc | Belgium | 119 |
| Bakirkoy Education and Research Hospital for Psychiatric and Neurological Diseases | Turkey | 112 |
| University of Florence | Italy | 110 |
| St. Michael's Hospital | Canada | 108 |
| Fakultni nemocnice Ostrava | Czechia | 106 |
| Neuro Rive-Sud | Canada | 102 |
| Fakultni nemocnice u sv. Anny v Brne | Czechia | 89 |
| Wexham Park Hospital, Frimley Health Foundation Trust | UK | 77 |
| Rehabilitation and MS-Centre Overpelt | Belgium | 71 |
| Zuyderland Medical Center | Netherlands (the) | 68 |
| Univ G. D’Annuzio Chieti-Pescara | Italy | 66 |
| Royal Hobart Hospital | Australia | 50 |
| Royal Brisbane and Women's Hospital | Australia | 49 |
| Monash Medical Centre | Australia | 46 |
| Ospedale Generale Provinciale Macerata | Italy | 45 |
| Hospital Universitario Nacional de Colombia | Colombia | 44 |
| Liverpool Hospital | Australia | 43 |
| Austin Health | Australia | 42 |
| Antwerp University Hospital | Belgium | 36 |
| Westmead Hospital | Australia | 35 |
| Universita di Foggia | Italy | 34 |
| Mayis University, Medical Faculty | Turkey | 33 |
| St Vincent's Hospital, Sydney | Australia | 29 |
| Nemocnice Ceske Budejovice, a.s. | Czechia | 29 |
| Centre Hospitalier Universitaire de Sherbrooke | Canada | 26 |

| | | |
|---|---|---|
| Brain and Mind Centre (BMRI) | Australia | 24 |
| Flinders Medical Centre | Australia | 24 |
| Fakultni nemocnice Kralovske Vinohrady | Czechia | 24 |
| Sultan Qaboos University Hospital | Oman | 24 |
| São João University Hospital | Portugal | 24 |
| University of Debrecen | Hungary | 23 |
| Razi hospital | Tunisia | 23 |
| West-Tallinn Central Hospital | Estonia | 22 |
| Centro Hospitalar Universitario de Sao Joao | Portugal | 18 |
| Hospital General Universitario de Alicante | Spain | 17 |
| University Medical Centre Ljubljana | Slovenia | 14 |
| AZ Alma | Belgium | 13 |
| Concord Repatriation General Hospital | Australia | 11 |
| Universitatsspital Basel | Switzerland | 11 |
| Mater Dei Hospital | Malta | 11 |
| Hospital Donostia | Spain | 10 |
| Az Sint-Jan Brugge | Belgium | 9 |
| Jewish General Hospital | Canada | 8 |
| Hospital Universitario de la Ribera | Spain | 8 |
| AHEPA University Hospital | Greece | 7 |
| Kyushu University Hospital | Japan | 7 |
| Fakultni Thomayerova nemocnice | Czechia | 6 |
| Tokyo Metropolitan Health and Medical Treatment Corporation Ebara Hospital | Japan | 6 |
| Nagoya City University School of Medical Sciences | Japan | 6 |
| Yamaguchi University Graduate School of Medicine | Japan | 6 |
| Koc University School of Medicine | Turkey | 6 |
| King Fahad Specialist Hospital-Dammam | Saudi Arabia | 5 |
| Ipswich hospital | UK | 4 |
| Hospital Germans Trias i Pujol | Spain | 3 |
| Veszprém Megyei Csolnoky Ferenc Kórház | Hungary | 3 |
| University Hospital Geelong | Australia | 2 |
| St Vincents Hospital, Fitzroy | Australia | 2 |
| The Townsville Hospital | Australia | 2 |
| CSSS Saint-Jérôme | Canada | 2 |
| St. Marianna Univertisy School of Medicine | Japan | 2 |
| Hospital Angeles de las Lomas. Instituto Mexicano de Neurociencias. | Mexico | 2 |
| Royal Hospital | Oman | 2 |
| Emergency Clinical County Hospital, Pius Brinzeu | Romania | 2 |
| Hacettepe University | Turkey | 2 |
| Haydarpasa Numune Training and Research Hospital | Turkey | 2 |
| Royal North Shore Hospital | Australia | 1 |
| Semmelweis University Budapest | Hungary | 1 |
| Péterfy Sandor Hospital | Hungary | 1 |
| BAZ County Hospital | Hungary | 1 |
| Szent Borbála Kórház | Hungary | 1 |
| Fukuoka Central Hospital | Japan | 1 |
| Ankara University Ibni Sina Hospital | Turkey | 1 |

**Supplementary Table 3. Characteristics of participants included in prognosis analyses**

| | Relapse analysis | | | Confirmed disability worsening analysis | | |
|---|---|---|---|---|---|---|
| | No CSL (*n*=8301) | CSL (*n*=1931) | SMD | No CSL (*n*=7175) | CSL (*n*=1726) | SMD |
| **Sex, number (%)** | | | 0.030 | | | 0.028 |
| **Female** | 5972 (71.9%) | 1363 (70.6%) | | 5168 (72.0%) | 1221 (70.7%) | |
| **Male** | 2329 (28.1%) | 568 (29.4%) | | 2007 (28.0%) | 505 (29.3%) | |
| **Age, mean (SD)** | 41.79 (10.56) | 38.79 (10.28) | 0.288 | 41.67 (10.48) | 38.67 (10.19) | 0.291 |
| **Disease duration, median (IQR)** | 8.29 (4.28, 14.47) | 7.04 (3.84, 12.02) | 0.206 | 8.18 (4.25, 14.31) | 6.98 (3.83, 12.02) | 0.196 |
| **Year, median (IQR)** | 2019 (2016, 2022) | 2017 (2014, 2020) | 0.491 | 2019 (2016, 2021) | 2017 (2014, 2020) | 0.474 |
| **EDSS score, median (IQR)** | 1.5 (1.0, 3.0) | 1.5 (1.0, 2.5) | 0.196 | 1.5 (1.0, 3.0) | 1.5 (1.0, 2.5) | 0.201 |
| **Number of relapses in previous 2 years, mean (SD)** | 0.24 (0.54) | 0.33 (0.61) | 0.150 | 0.26 (0.55) | 0.35 (0.62) | 0.148 |
| **Country of care, number (%)** | | | 0.544 | | | 0.539 |
| **Czechia** | 1768 (21.3%) | 387 (20.0%) | | 1659 (23.1%) | 369 (21.4%) | |
| **Australia** | 1626 (19.6%) | 181 (9.4%) | | 1414 (19.7%) | 160 (9.3%) | |
| **Italy** | 1121 (13.5%) | 599 (31.0%) | | 977 (13.6%) | 551 (31.9%) | |
| **Turkey** | 1007 (12.1%) | 224 (11.6%) | | 868 (12.1%) | 199 (11.5%) | |
| **Spain** | 578 (7.0%) | 169 (8.8%) | | 533 (7.4%) | 159 (9.2%) | |
| **Canada** | 495 (6.0%) | 98 (5.1%) | | 412 (5.7%) | 77 (4.5%) | |
| **Belgium** | 366 (4.4%) | 60 (3.1%) | | 316 (4.4%) | 49 (2.8%) | |
| **GB** | 368 (4.4%) | 30 (1.6%) | | 295 (4.1%) | 27 (1.6%) | |
| **Lebanon** | 299 (3.6%) | 52 (2.7%) | | 271 (3.8%) | 47 (2.7%) | |
| **Croatia** | 223 (2.7%) | 23 (1.2%) | | 83 (1.2%) | 10 (0.6%) | |
| **Kuwait** | 191 (2.3%) | 32 (1.7%) | | 178 (2.5%) | 31 (1.8%) | |
| **Netherlands** | 55 (0.7%) | 13 (0.7%) | | 45 (0.6%) | 13 (0.8%) | |
| **Colombia** | 41 (0.5%) | 3 (0.2%) | | 0 (0.0%) | 0 (0.0%) | |
| **Portugal** | 25 (0.3%) | 17 (0.9%) | | 0 (0.0%) | 0 (0.0%) | |
| **Other** | 138 (1.7%) | 43 (2.2%) | | 124 (1.7%) | 34 (2.0%) | |
| **Duration of baseline DMT, years, median (IQR)** | 2.57 [1.90, 4.18] | 2.60 [1.92, 4.25] | 0.004 | 2.54 (1.89, 4.06) | 2.55 (1.91, 4.28) | 0.030 |
| **DMT efficacy group, number (%)** | | | 0.617 | | | 0.615 |
| **Platform** | 3301 (39.8%) | 1247 (64.6%) | | 2970 (41.4%) | 1142 (66.2%) | |
| **Moderate efficacy** | 2417 (29.1%) | 497 (25.7%) | | 2033 (28.3%) | 426 (24.7%) | |
| **High efficacy** | 2583 (31.1%) | 187 (9.7%) | | 2172 (30.3%) | 158 (9.2%) | |
| **Number of CSLs on index MRI, number (%)** | | | NA | | | NA |
| **1** | NA | 939 (48.6%) | | NA | 835 (48.4%) | |
| **2** | NA | 369 (19.1%) | | NA | 334 (19.4%) | |
| **3** | NA | 210 (10.9%) | | NA | 188 (10.9%) | |
| **4 to 9** | NA | 275 (14.2%) | | NA | 240 (13.9%) | |
| **10 or more** | NA | 138 (7.1%) | | NA | 129 (7.5%) | |
| **Contrast administered for index MRI, number (%)** | 2960 (35.7%) | 978 (50.6%) | NA | 2630 (36.7%) | 893 (51.7%) | NA |
| **Only non-enhancing lesions, number (%)** | NA | 575 (29.8%) | | NA | 535 (31.0%) | |
| **Any contrast-enhancing lesions, number (%)** | NA | 403 (20.9%) | | NA | 358 (20.7%) | |
| **Follow-up duration, years, median (IQR)** | 3.83 (1.77, 6.35) | 4.80 (2.36, 7.45) | 0.248 | 4.36 (2.54, 6.78) | 5.31 (3.05, 7.68) | 0.241 |
| **Follow-up clinic visits per year, median (IQR)** | 2.00 (1.47, 2.59) | 2.08 (1.64, 2.79) | 0.118 | 2.06 (1.60, 2.67) | 2.16 (1.72, 2.83) | 0.094 |

All listed characteristics are at baseline except follow-up duration and follow-up clinic visits per year. Abbreviations: CSL, clinically silent lesion; DMT, disease modifying therapy; EDSS, expanded disability status scale; IQR, interquartile range; SD, standard deviation; SMD, standardised mean difference, quantified using Cohen d value.

**Supplementary Table 4. Baseline DMTs for participants included in prognosis analyses**

| DMT | | Relapse analysis, number (%) | Confirmed disability worsening analysis, number (%) |
|---|---|---|---|
| **Platform DMTs** | **Interferon-beta** | 2591 (25.3%) | 2401 (27.0%) |
| | **Glatiramer acetate** | 1184 (11.6%) | 1068 (12.0%) |
| | **Teriflunomide** | 773 (7.6%) | 643 (7.2%) |
| **Moderate-efficacy DMTs** | **Fingolimod** | 1640 (16.0%) | 1469 (16.5%) |
| | **Dimethyl fumarate** | 997 (9.7%) | 820 (9.2%) |
| | **Cladribine** | 259 (2.5%) | 166 (1.9%) |
| | **Ozanimod** | 9 (0.1%) | 3 (0.0%) |
| | **Ponesimod** | 5 (0.0%) | 0 (0.0%) |
| | **Siponimod** | 3 (0.0%) | 0 (0.0%) |
| | **Diroximel fumarate** | 1 (0.0%) | 1 (0.0%) |
| **High-efficacy DMTs** | **Natalizumab** | 1466 (14.3%) | 1317 (14.8%) |
| | **Ocrelizumab** | 809 (7.9%) | 640 (7.2%) |
| | **Alemtuzumab** | 298 (2.9%) | 245 (2.8%) |
| | **Ofatumumab** | 77 (0.8%) | 25 (0.3%) |
| | **HSCT** | 64 (0.6%) | 54 (0.6%) |
| | **Rituximab** | 56 (0.5%) | 49 (0.6%) |

Abbreviations: DMT, disease modifying therapy; HSCT, haematopoietic stem cell transplantation

**Supplementary Table 5. Unadjusted and adjusted hazard ratios from Cox models for relapse and CDW after the index MRI**

| | Relapse analysis | | Confirmed disability worsening analysis | |
|---|---|---|---|---|
| | Unadjusted HR (95% CI) | Adjusted HR (95% CI) | Unadjusted HR (95% CI) | Adjusted HR (95% CI) |
| CSL vs. no CSL | 2.20 (1.98-2.45) | 1.76 (1.57-1.97) | 1.23 (1.06-1.43) | 1.38 (1.18-1.62) |
| Male vs. female | 0.84 (0.75-0.94) | 0.82 (0.73-0.92) | 0.96 (0.83-1.10) | 0.98 (0.86-1.13) |
| Age (years) | 0.98 (0.97-0.98) | 0.98 (0.97-0.98) | 1.02 (1.02-1.03) | 1.02 (1.02-1.03) |
| Age^2 | 1.00 (1.00-1.00) | 1.00 (1.00-1.00) | 1.00 (1.00-1.00) | 1.00 (1.00-1.00) |
| Calendar year | 0.93 (0.92-0.95) | 0.94 (0.88-0.99) | 0.97 (0.96-0.99) | 1.05 (0.96-1.14) |
| Calendar year^2 | 1.00 (1.00-1.00) | 1.00 (1.00-1.00) | 1.00 (1.00-1.00) | 1.00 (0.99-1.00) |
| Disease duration (years) | 0.98 (0.97-0.99) | 1.01 (0.99-1.04) | 1.02 (1.01-1.03) | 1.00 (0.98-1.03) |
| Disease duration^2 | 1.00 (1.00-1.00) | 1.00 (1.00-1.00) | 1.00 (1.00-1.00) | 1.00 (1.00-1.00) |
| Baseline EDSS score | 1.05 (1.02-1.08) | 1.05 (0.86-1.27) | 1.12 (1.08-1.16) | 0.91 (0.73-1.14) |
| Baseline EDSS score^2 | 1.00 (1.00-1.01) | 1.07 (0.99-1.16) | 1.02 (1.01-1.02) | 1.07 (0.98-1.17) |
| Baseline EDSS score^3 | 1.00 (1.00-1.00) | 0.99 (0.98-1.00) | 1.00 (1.00-1.00) | 0.99 (0.98-1.00) |
| Moderate vs. low efficacy DMT | 0.74 (0.66-0.84) | 0.92 (0.81-1.06) | 1.12 (0.96-1.30) | 1.05 (0.89-1.25) |
| High vs. low efficacy DMT | 0.52 (0.46-0.60) | 0.65 (0.55-0.76) | 1.33 (1.15-1.54) | 1.19 (0.99-1.43) |
| Number of relapses in the last 2 years | 1.62 (1.52-1.73) | 1.44 (1.34-1.55) | 1.02 (0.92-1.14) | 1.07 (0.96-1.19) |

Hazard ratios are at 2 years of follow up. Abbreviations: CSL, clinically silent lesion; DMT, disease modifying therapy; EDSS, expanded disability status scale.

**Supplementary Table 6. Estimated hazard ratios for alternative CSL comparisons and subgroup analyses comparing CSLs to no CSLs, from Cox models for relapse and CDW after the index MRI**

| | | Relapse | | Confirmed disability worsening | |
|---|---|---|---|---|---|
| | | Unadjusted HR (95% CI) | Adjusted HR (95% CI) | Unadjusted HR (95% CI) | Adjusted HR (95% CI) |
| **Alternative CSL comparisons** | | | | | |
| **Number of CSLs** | **1 CSL** | 1.95 (1.68, 2.25) | 1.59 (1.37, 1.85) | 1.17 (0.95, 1.43) | 1.35 (1.09, 1.66) |
| | **≥2 CSLs** | 2.46 (2.15, 2.81) | 1.94 (1.68, 2.24) | 1.29 (1.07, 1.56) | 1.42 (1.15, 1.74) |
| **Contrast enhancement** | **CEL** | 2.36 (1.92, 2.90) | 1.79 (1.44, 2.22) | 1.04 (0.76, 1.43) | 1.17 (0.84, 1.63) |
| | **Non-CEL** | 1.80 (1.48, 2.19) | 1.53 (1.25, 1.88) | 1.24 (0.96, 1.59) | 1.34 (1.03, 1.75) |
| **Subgroup analyses** | | | | | |
| **DMT efficacy** | **Platform** | 1.86 (1.62, 2.14) | 1.66 (1.44, 1.92) | 1.30 (1.06, 1.59) | 1.29 (1.04, 1.59) |
| | **Moderate-efficacy** | 2.45 (1.98, 3.02) | 2.14 (1.71, 2.68) | 1.37 (1.03, 1.83) | 1.49 (1.11, 2.01) |
| | **High-efficacy** | 1.80 (1.22, 2.64) | 1.36 (0.90, 2.04) | 1.41 (0.96, 2.09) | 1.64 (1.08, 2.48) |
| **Disease duration subgroups** | **0-5 years** | 2.13 (1.78, 2.55) | 1.65 (1.37, 2.00) | 1.15 (0.87, 1.51) | 1.25 (0.93, 1.67) |
| | **5-10 years** | 2.00 (1.64, 2.43) | 1.55 (1.26, 1.91) | 1.26 (0.96, 1.66) | 1.42 (1.05, 1.90) |
| | **≥10 years** | 2.41 (2.01, 2.90) | 1.99 (2.63, 2.44) | 1.33 (1.06, 1.67) | 1.46 (1.14, 1.88) |

Hazard ratios are at 2 years of follow up. Abbreviations: CEL, contrast enhancing lesion; CSL, clinically silent lesion; DMT, disease modifying therapy.

**Supplementary Table 7. Adjusted hazard ratios and 95% CIs for sensitivity analyses comparing CSLs to no CSLs from Cox models for relapse and disability worsening after the index MRI**

| | Relapse | Disability worsening[a] |
|---|---|---|
| **Adjusted for country health expenditure[b]** | 1.78 (1.59-1.99) | 1.39 (1.19-1.63) |
| **Among participants with available data on the total lesion count[c]** | | |
| **Total lesion count included in models** | 1.48 (1.18-1.85) | 1.30 (0.98-1.72) |
| **Total lesion count not included in models** | 1.49 (1.20-1.86) | 1.32 (1.00, 1.75) |
| **Unconfirmed disability worsening[d]** | NA | 1.39 (1.22-1.58) |

Adjusted hazard ratios (95% CIs) are at 2 years of follow up. [a]Disability worsening outcomes use the 6-month confirmed disability worsening (CDW) definition for all analyses in this column except the unconfirmed disability worsening row, which is defined as an EDSS score increase as per the CDW definition, but without the requirement for later confirmation of the EDSS score increase. [b]Cox models without country stratification and with adjustment for health expenditure in the country of care (categorised according to World Health Organisation global health expenditure data, 2016[5] as <1500, 1500-3000 or ≥3000 US dollars per capita per annum).[c]Among participants with available data on the total lesion count on their index MRI scan, relapses occurred in 352 (22.3%) of 1,579 participants without CSLs and 262 (38.0%) of 689 participants with CSLs, and CDW occurred in 299 (21.6%) of 1,386 participants without CSLs and 182 (28.5%) of 639 participants with CSLs. [d]Unconfirmed disability worsening occurred in 1,225 (17.1%) of 7,175 participants without CSLs and 351 (20.3%) of 1,726 participants with CSLs. Abbreviations: CSL, clinically silent lesion.

**Supplementary Table 8. Country of care of participants included in the emulated trials of DMT escalation after clinically silent lesions**

| | Relapse analysis | | | Confirmed disability worsening analysis | | |
|---|---|---|---|---|---|---|
| | **All participants[a] (*n*=2264)** | **Participants who escalated DMT within 6 months (*n*=286)** | **SMD** | **All participants[a] (*n*=2016)** | **Participants who escalated DMT within 6 months (*n*=248)** | **SMD** |
| **Italy** | 731 (32.3%) | 34 (11.9%) | 0.693 | 675 (33.5%) | 33 (13.3%) | 0.696 |
| **Czechia** | 496 (21.9%) | 42 (14.7%) | | 467 (23.2%) | 38 (15.3%) | |
| **Turkey** | 264 (11.7%) | 37 (12.9%) | | 239 (11.9%) | 35 (14.1%) | |
| **Australia** | 207 (9.1%) | 76 (26.6%) | | 182 (9.0%) | 69 (27.8%) | |
| **Spain** | 188 (8.3%) | 34 (11.9%) | | 174 (8.6%) | 29 (11.7%) | |
| **Canada** | 109 (4.8%) | 12 (4.2%) | | 84 (4.2%) | 10 (4.0%) | |
| **Lebanon** | 66 (2.9%) | 15 (5.2%) | | 63 (3.1%) | 15 (6.0%) | |
| **Belgium** | 54 (2.4%) | 8 (2.8%) | | 47 (2.3%) | 8 (3.2%) | |
| **Other** | 149 (6.6%) | 28 (9.8%) | | 85 (4.2%) | 11 (4.4%) | |

Emulated trial outcome models were adjusted for country of care. [a]All but 0.4% participants (10 of 2264 for the relapse analysis and 9 of 2016 for the CDW analysis) of were included in the control arm for at least the first week's emulated trial. Abbreviations: DMT, disease modifying therapy; SMD, standardised mean difference, quantified using Cohen d value.

**Supplementary Table 9. Baseline DMTs and DMT group changes during follow-up for participants included in the emulated trials**

| | | Relapse analysis (*n*=2,264) | CDW analysis (*n*=2,016) |
|---|---|---|---|
| **Baseline DMTs** | **Platform DMTs** | 1585 (70.0%) | 1,431 (71.0%) |
| | **Interferon-beta** | 915 (40.4%) | 856 (42.5%) |
| | **Glatiramer acetate** | 423 (18.7%) | 371 (18.4%) |
| | **Teriflunomide** | 247 (10.9%) | 204 (10.1%) |
| | **Moderate-efficacy DMTs** | 679 (30.0%) | 585 (29.0%) |
| | **Fingolimod** | 393 (17.4%) | 346 (17.2%) |
| | **Dimethyl fumarate** | 230 (10.2%) | 205 (10.2%) |
| | **Cladribine** | 56 (2.5%) | 34 (1.7%) |
| **DMT group changes during follow-up**[a] | **Unchanged DMT efficacy**[b] | 1,922 (85.0%) | 1,703 (84.5%) |
| | **Changed to a DMT of equivalent efficacy**[b] | 56 (2.5%) | 65 (3.2%) |
| | **Escalated DMT**[c] | 286 (12.6%) | 248 (12.3%) |
| | **Platform baseline DMT escalated to moderate-efficacy DMT** | 153 (6.8%) | 135 (6.7%) |
| | **Platform baseline DMT escalated to high-efficacy DMT** | 45 (2.0%) | 36 (1.8%) |
| | **Moderate-efficacy baseline DMT escalated to high-efficacy DMT** | 88 (3.9%) | 77 (3.8%) |

[a]DMT group changes are during the 6-month treatment allocation window after CSLs. DMT changes are included if they occurred before another competing outcome or censoring event. [b]These participants remained in the control arm for all trial weeks until they were censored. [c]These participants were censored from the control arm after they escalated DMT and then included in the DMT escalation arm for all subsequent trial weeks until they were censored. Abbreviations: CDW, confirmed disability worsening; DMT, disease-modifying therapy.

**Supplementary Table 10. Adjusted hazard ratios (95% confidence intervals) from emulated trial outcome models**

| | Relapse | Confirmed disability worsening |
|---|---|---|
| **DMT escalation vs. control** | 0.34 (0.23-0.47) | 0.89 (0.56-1.33) |
| **Male vs. female** | 0.86 (0.71-1.06) | 1.06 (0.77-1.43) |
| **Age (years)** | 0.97 (0.95-0.98) | 1.02 (1.00-1.03) |
| **Age^2** | 1.00 (1.00-1.00) | 1.00 (1.00-1.00) |
| **Calendar year** | 0.92 (0.84-1.01) | 1.11 (0.96-1.29) |
| **Calendar year^2** | 1.00 (1.00-1.01) | 0.99 (0.98-1.00) |
| **Disease duration** | 1.01 (0.96-1.06) | 1.03 (0.98-1.10) |
| **Disease duration^2** | 1.00 (1.00-1.00) | 1.00 (1.00-1.00) |
| **EDSS score** | 0.87 (0.61-1.24) | 1.18 (0.67-2.01) |
| **EDSS score^2** | 1.18 (1.01-1.41) | 1.00 (0.81-1.28) |
| **EDSS score^3** | 0.98 (0.96-1.00) | 1.00 (0.97-1.02) |
| **Moderate vs. low efficacy DMT** | 0.89 (0.69-1.11) | 1.07 (0.70-1.56) |
| **Number of relapses in the previous 2 years** | 1.48 (1.28-1.69) | 1.15 (0.87-1.47) |
| **Country of care (vs. Australia)** | | |
| **Czechia** | 1.23 (0.80-2.15) | 0.76 (0.41-1.69) |
| **Italy** | 0.58 (0.37-0.97) | 0.82 (0.41-1.64) |
| **Turkey** | 0.89 (0.54-1.56) | 0.83 (0.36-1.79) |
| **Spain** | 0.85 (0.49-1.51) | 0.86 (0.41-1.86) |
| **Canada** | 0.80 (0.44-1.55) | 0.80 (0.28-2.02) |
| **Belgium** | 0.60 (0.21-1.33) | 1.83 (0.55-4.35) |
| **Lebanon** | 0.44 (0.16-0.93) | 0.84 (0.22-2.19) |
| **Other** | 0.82 (0.46-1.58) | 1.27 (0.55-2.86) |
| **Number of CSLs (vs. 1)** | | |
| **2** | 1.27 (1.01-1.63) | 0.90 (0.59-1.29) |
| **3** | 1.37 (0.98-1.86) | 1.38 (0.78-2.30) |
| **4 to 9** | 1.25 (0.89-1.72) | 0.93 (0.58-1.41) |
| **10 or more** | 1.28 (0.86-1.93) | 1.11 (0.63-1.88) |
| **Contrast enhancement (vs non-enhancing)** | | |
| **Enhancing** | 1.15 (0.87-1.53) | 0.82 (0.48-1.27) |
| **Contrast not administered** | 0.95 (0.73-1.21) | 0.97 (0.68-1.35) |
| **Weeks post MRI** | 0.99 (0.98-0.99) | 0.98 (0.98-0.99) |
| **Weeks post MRI^2** | 1.00 (1.00-1.00) | 1.00 (1.00-1.00) |

Abbreviations: CSL, clinically silent lesion; DMT, disease modifying therapy; EDSS, expanded disability status scale.

**Supplementary Table 11. Censoring weight metrics from emulated trials**

| | Relapse | | Confirmed disability worsening | |
|---|---|---|---|---|
| | **Median (IQR)** | **Range** | **Median (IQR)** | **Range** |
| **Censoring weight 1[a]** | | | | |
| **Unstabilised** | 1.07 (1.03-1.61) | 1.00-19.82 | 1.07 (1.03-1.15) | 1.00-21.97 |
| **Stabilised** | 0.98 (0.92-1.00) | 0.64-13.11 | 0.98 (0.93-1.00) | 0.65-15.78 |
| **Censoring weight 2[b]** | | | | |
| **Unstabilised** | 1.00 (1.00-1.02) | 1.00-8.30 | 1.00 (1.00-1.00) | 1.00-10.33 |
| **Stabilised** | 1.00 (1.00-1.00) | 0.37-3.11 | 1.00 (1.00-1.00) | 0.42-4.37 |
| **Final censoring weight[c]** | 0.97 (0.92-1.01) | 0.33-13.11 | 0.97 (0.92-1.00) | 0.418-17.87 |

[a]Censoring from the control group because of DMT escalation despite no clinical event. [b]Censoring from the control group because of no DMT escalation despite a clinical event. [c]The product of censoring weight 1 and censoring weight 2.

**Supplementary Table 12. Adjusted hazard ratios and 95% CIs for sensitivity analyses comparing DMT escalation and control treatment strategies from emulated trials**

| | **Relapse** | **Disability worsening[a]** |
|---|---|---|
| **Adjusted for country health expenditure[b]** | 0.34 (0.23-0.47) | 0.93 (0.61-1.31) |
| **Unconfirmed disability worsening[c]** | NA | 0.84 (0.59-1.15) |
| **Alternative control treatment strategies** | | |
| **(a) No requirement for DMT escalation following clinical events** | 0.36 (0.25-0.50) | 0.83 (0.54-1.21) |
| **(b) Additionally, no requirement for unchanged DMT unless a clinical event occurs** | 0.44 (0.31-0.59) | 0.84 (0.58-1.15) |

Adjusted hazard ratios (95% CIs) are at 2 years of follow up. [a]Disability worsening outcomes use the 6-month confirmed disability worsening (CDW) definition for all analyses in this column except the unconfirmed disability worsening row, which is defined as an EDSS score increase as per the CDW definition, but without the requirement for later confirmation of the EDSS score increase. [b]Outcome and censoring models adjusted for health expenditure in the country of care (categorised according to World Health Organisation global health expenditure data, 2016[5] as <1500, 1500-3000 or ≥3000 US dollars per capita per annum) instead of a marker of the country. Abbreviations: DMT, disease-modifying therapy.

### Supplementary figures

#### Supplementary Figure 1. Summary of alternative CSL comparisons and subgroup analyses comparing the probability of outcomes in participants with and without CSLs

##### A. Relapse

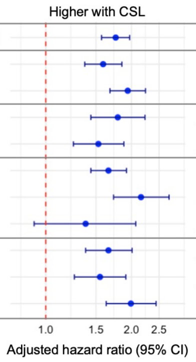

| | | No CSL n | CSL n | No CSL cumulative incidence (95% CI) | CSL cumulative incidence (95% CI) | aHR [95% CI] | Heterogeneity[a] |
|---|---|---|---|---|---|---|---|
| | ALL PARTICIPANTS | 8301 | 1931 | 14.3% [13.5%, 15.2%] | 27.8% [25.7%, 29.9%] | 1.76 [1.57, 1.97] | |
| Number of CSLs | 1 CSL | 8301 | 939 | 14.3% [13.5%, 15.2%] | 25.1% [22.3%, 28.2%] | 1.59 [1.37, 1.85] | p=0.06 |
| | ≥2 CSLs | 8301 | 992 | 14.3% [13.5%, 15.2%] | 30.3% [27.4%, 33.4%] | 1.94 [1.68, 2.24] | |
| Contrast enhancement | CEL | 2960 | 403 | 15.5% [14.1%, 16.9%] | 30.8% [26.4%, 35.8%] | 1.79 [1.44, 2.22] | p=0.30 |
| | Non-CEL | 2960 | 575 | 15.5% [14.1%, 16.9%] | 25.4% [21.9%, 29.4%] | 1.53 [1.25, 1.88] | |
| DMT efficacy | Platform | 3301 | 1247 | 17.4% [16.1%, 18.9%] | 28.9% [26.4%, 31.6%] | 1.66 [1.44, 1.92] | p=0.08 |
| | Moderate-efficacy | 2417 | 497 | 13.3% [11.8%, 14.8%] | 28.2% [24.3%, 32.7%] | 2.14 [1.71, 2.68] | |
| | High-efficacy | 2583 | 187 | 11.0% [9.8%, 12.4%] | 18.2% [13.0%, 25.2%] | 1.36 [0.90, 2.04] | |
| Disease duration | 0-5 years | 2511 | 678 | 15.8% [14.2%, 17.3%] | 29.2% [25.6%, 32.7%] | 1.65 [1.37, 2.00] | p=0.21 |
| | 5-10 years | 2323 | 605 | 15.0% [13.4%, 16.6%] | 26.9% [23.1%, 30.5%] | 1.55 [1.26, 1.91] | |
| | ≥10 years | 3467 | 648 | 12.8% [11.6%, 14.0%] | 27.0% [23.3%, 30.5%] | 1.99 [2.63, 2.44] | |

##### B. Confirmed disability worsening

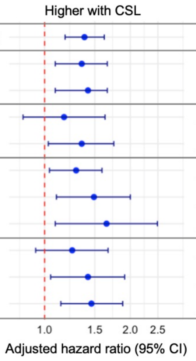

| | | No CSL n | CSL n | No CSL cumulative incidence (95% CI) | CSL cumulative incidence (95% CI) | aHR [95% CI] | Heterogeneity[a] |
|---|---|---|---|---|---|---|---|
| | ALL PARTICIPANTS | 7175 | 1726 | 11.4% [10.7%, 12.2%] | 13.8% [12.2%, 15.5%] | 1.38 [1.18, 1.62] | |
| Number of CSLs | 1 CSL | 7175 | 835 | 11.4% [10.7%, 12.2%] | 13.3% [11.1%, 15.8%] | 1.35 [1.09, 1.66] | p=0.74 |
| | ≥2 CSLs | 7175 | 891 | 11.4% [10.7%, 12.2%] | 14.2% [12.1%, 16.8%] | 1.42 [1.15, 1.74] | |
| Contrast enhancement | CEL | 2630 | 358 | 12.4% [11.1%, 13.7%] | 12.7% [9.6%, 16.6%] | 1.17 [0.84, 1.63] | p=0.51 |
| | Non-CEL | 2630 | 535 | 12.4% [11.1%, 13.7%] | 15.0% [12.1%, 18.3%] | 1.34 [1.03, 1.75] | |
| DMT efficacy | Platform | 2970 | 1142 | 10.0% [9.0%, 11.2%] | 12.7% [10.8%, 14.8%] | 1.29 [1.04, 1.59] | p=0.51 |
| | Moderate-efficacy | 2033 | 426 | 11.3% [10.0%, 12.8%] | 14.8% [11.7%, 18.7%] | 1.49 [1.11, 2.01] | |
| | High-efficacy | 2172 | 158 | 13.4% [12.0%, 15.0%] | 19.0% [13.5%, 26.4%] | 1.64 [1.08, 2.48] | |
| Disease duration | 0-5 years | 2185 | 610 | 10.3% [9.0%, 11.6%] | 11.6% [8.9%, 14.1%] | 1.25 [0.93, 1.67] | p=0.72 |
| | 5-10 years | 2037 | 538 | 10.8% [9.4%, 12.2%] | 13.2% [10.3%, 16.1%] | 1.42 [1.05, 1.90] | |
| | ≥10 years | 2953 | 578 | 12.7% [11.4%, 13.9%] | 16.6% [13.4%, 19.6%] | 1.46 [1.14, 1.88] | |

Dots represent 2-year hazard ratios from Cox models adjusted for baseline confounders, and horizontal bars represent their 95% confidence intervals. Within each subgroup, aHRs compare outcomes in participants with CSLs to those without CSLs. Cumulative incidences are derived from unadjusted Kaplan-Meier estimates. [a]Heterogeneity between subgroup log hazard ratios was tested using Cochran's Q test. Abbreviations: aHR, adjusted hazard ratio; CEL, contrast-enhancing lesion; CSL, clinically silent lesion.

**Supplementary Figure 2. Inclusion flow diagram for analyses of confirmed disability worsening outcomes**

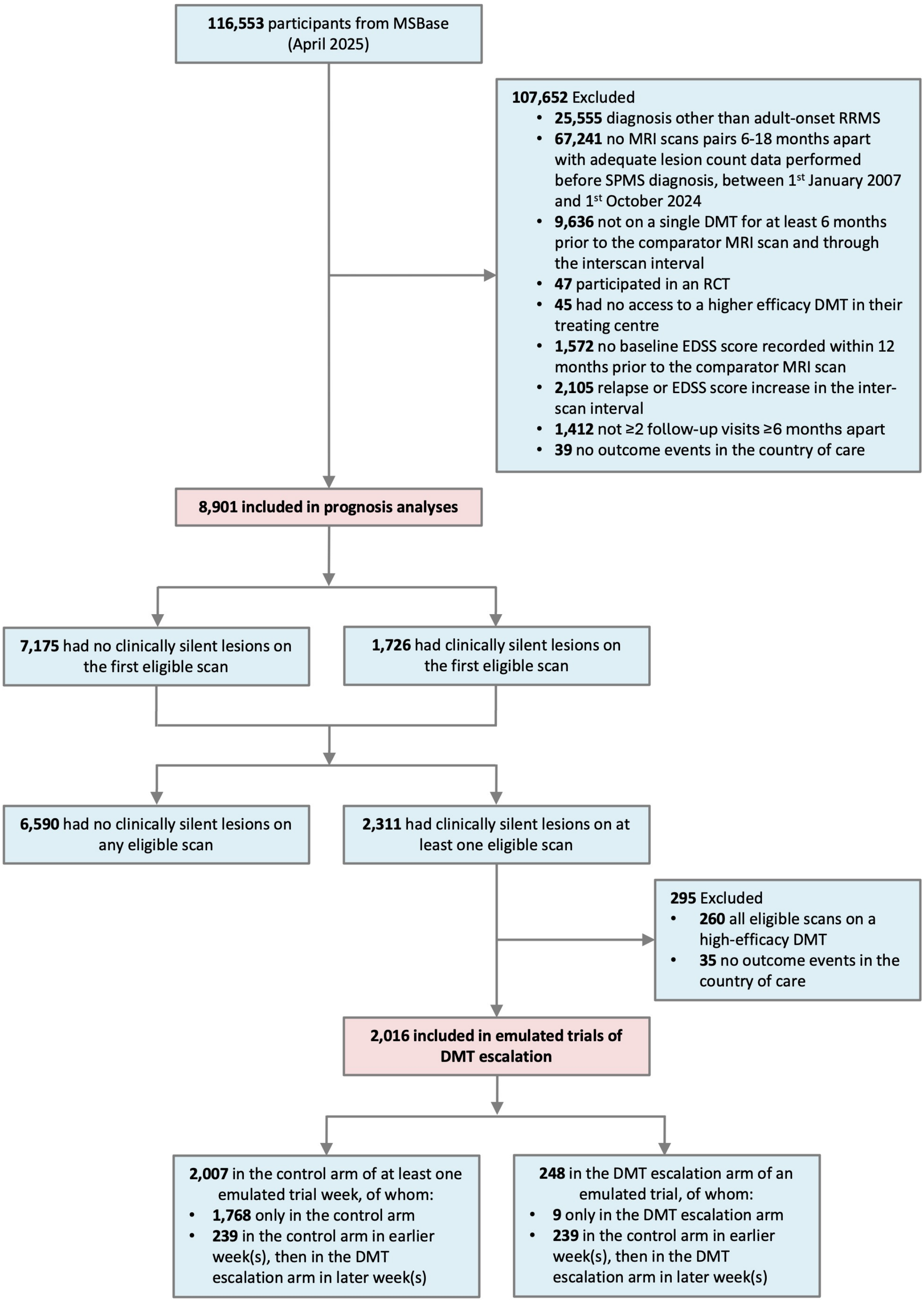

Abbreviations: DMT, disease modifying therapy; EDSS, Expanded Disability Status Scale; RCT, Randomised Controlled Trial; RRMS, relapsing-remitting multiple sclerosis; SPMS, secondary progressive multiple sclerosis.

**Supplementary Figure 3. Proportion of included participants who escalated DMT within 6 months following CSLs by calendar year epoch**

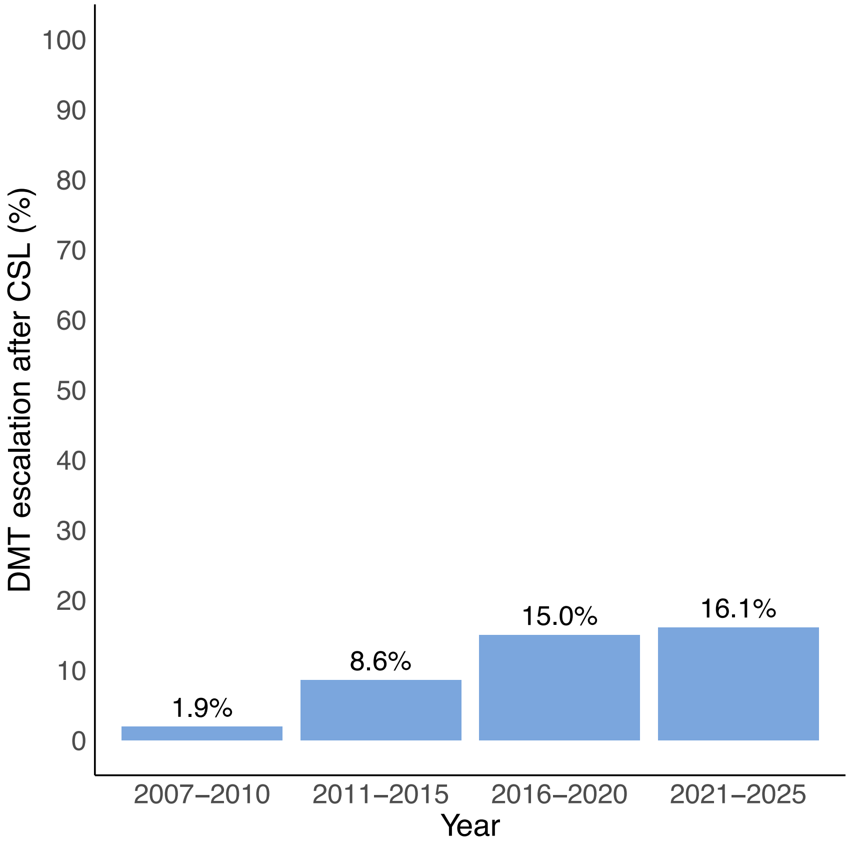

Abbreviations: CSL, clinically silent lesion. DMT, disease modifying therapy.

**Supplementary Figure 4. Stacked cumulative incidence curves of competing events in the emulated trial of the effect of DMT escalation following CSLs on relapses**

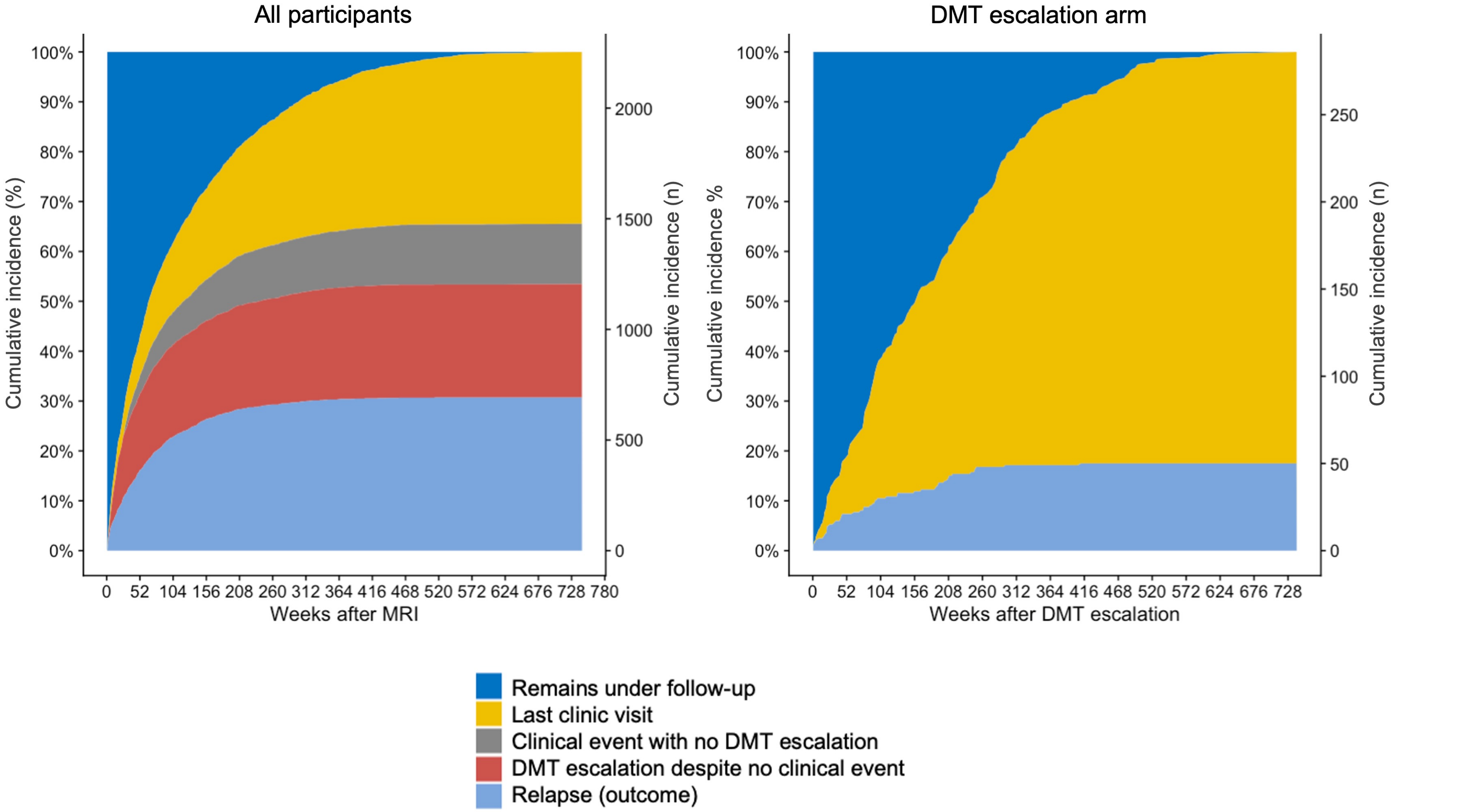

Stacked cumulative incidence curves plotting the first of the competing events during follow-up for participants in the emulated trial of DMT escalation after CSLs on the probability of subsequent relapses. Left panel shows all participants (n=2,285), starting from the time of their MRI scan. Right panel shows participants who were included in the DMT escalation arm of a trial (n=288), starting from the time of DMT escalation. Abbreviations: CSL, clinically silent lesion. DMT, disease modifying therapy.

**Supplementary Figure 5. Summary of subgroup analyses estimating the effect of DMT escalation after CSLs on the probability of relapses and CDW**

### A. Relapse

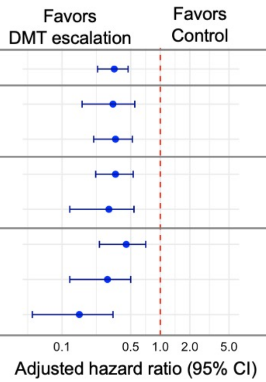

| | | Control cumulative incidence [95% CI] | DMT escalation cumulative incidence [95% CI] | aHR [95% CI] | Heterogeneity[a] |
|---|---|---|---|---|---|
| | ALL PARTICIPANTS | 38.9% [35.8%, 42.1%] | 16.8% [12.4%, 23.4%] | 0.34 [0.23, 0.47] | |
| Number of CSLs | 1 CSL | 36.5% [32.6%, 41.1%] | 15.1% [7.9%, 24.9%] | 0.33 [0.16, 0.55] | p=0.880 |
| | ≥2 CSLs | 41.9% [37.5%, 46.4%] | 18.8% [11.6%, 26.7%] | 0.35 [0.21, 0.52] | |
| DMT efficacy | Platform | 38.9% [35.4%, 42.5%] | 18.6% [12.0%, 27.9%] | 0.35 [0.22, 0.53] | p=0.729 |
| | Moderate-efficacy | 41.5% [35.3%, 48.8%] | 13.6% [6.4%, 22.0%] | 0.30 [0.12, 0.54] | |
| Disease duration | 0-5 years | 43.0% [37.3%, 48.5%] | 23.7% [14.3%, 34.4%] | 0.45 [0.24, 0.71] | p=0.131 |
| | 5-10 years | 40.3% [34.4%, 46.0%] | 13.9% [7.0%, 22.1%] | 0.29 [0.12, 0.50] | |
| | ≥10 years | 35.1% [30.2%, 40.0%] | 11.9% [2.7%, 30.9%] | 0.15 [0.05, 0.33] | |

### B. Confirmed disability worsening

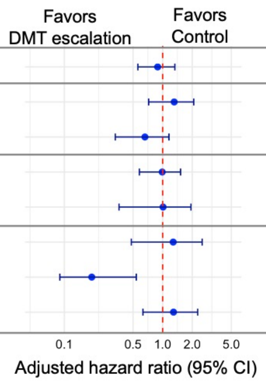

| | | Control cumulative incidence [95% CI] | DMT escalation cumulative incidence [95% CI] | aHR [95% CI] | Heterogeneity[a] |
|---|---|---|---|---|---|
| | ALL PARTICIPANTS | 17.7% [15.3%, 20.1%] | 16.0% [10.8%, 22.2%] | 0.89 [0.56, 1.33] | |
| Number of CSLs | 1 CSL | 17.7% [14.4%, 21.4%] | 21.6% [13.3%, 29.5%] | 1.31 [0.72, 2.07] | p=0.102 |
| | ≥2 CSLs | 17.5% [14.1%, 21.2%] | 12.6% [6.6%, 20.2%] | 0.66 [0.33, 1.16] | |
| DMT efficacy | Platform | 17.5% [14.7%, 20.3%] | 17.8% [11.0%, 25.3%] | 0.99 [0.58, 1.52] | p=0.968 |
| | Moderate-efficacy | 17.5% [13.3%, 22.0%] | 17.3% [7.3%, 27.2%] | 1.01 [0.36, 1.94] | |
| Disease duration | 0-5 years | 13.5% [9.9%, 17.9%] | 15.9% [6.6%, 25.9%] | 1.28 [0.48, 2.52] | p=0.001 |
| | 5-10 years | 18.3% [13.9%, 22.9%] | 3.9% [2.0%, 10.4%] | 0.19 [0.09, 0.54] | |
| | ≥10 years | 20.4% [15.9%, 24.8%] | 27.4% [15.1%, 41.3%] | 1.29 [0.63, 2.27] | |

Dots represent 4-year adjusted hazard ratios and horizontal bars represent their 95% confidence intervals. [a]Heterogeneity between subgroup log hazard ratios was tested using Cochran's Q test. Abbreviations: aHR, adjusted hazard ratio; CEL, contrast-enhancing lesion; CSL, clinically silent lesion.

**Supplementary Figure 6. Estimated effect of DMT escalation after CSLs on the probability of RAW and PIRA**

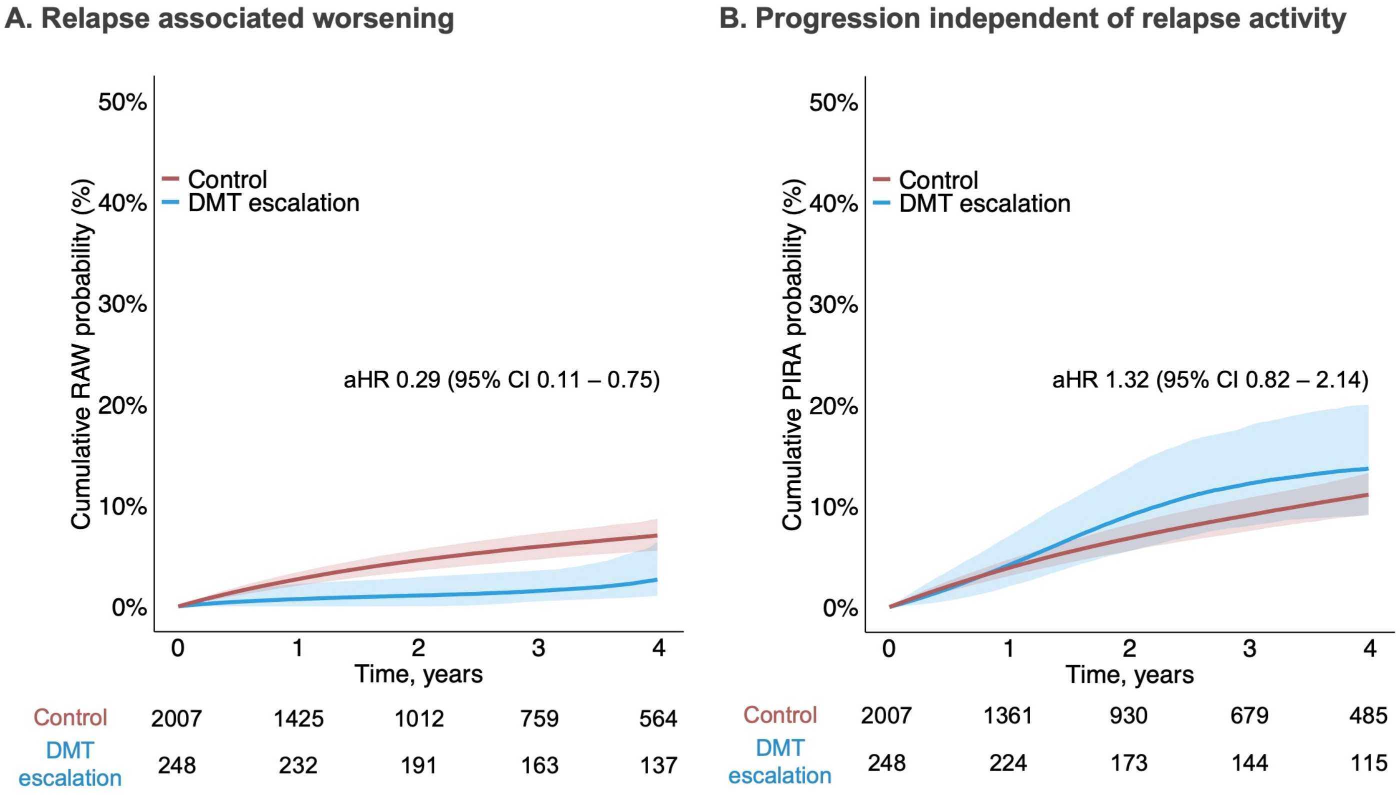

Cumulative probabilities were generated using G-computation; adjusted hazard ratios are from pooled logistic regression models for the emulated trial outcomes. Lines represent median probabilities and shaded areas represent 95% confidence intervals. Abbreviations: aHR, adjusted hazard ratio; DMT, disease modifying therapy; PIRA, progression independent of relapse activity; RAW, relapse-associated worsening.
